# DiabetIA: Building Machine Learning Models for Type 2 Diabetes Complications

**DOI:** 10.1101/2023.10.22.23297277

**Authors:** Joaquin Tripp, Daniel Santana-Quinteros, Rafael Perez-Estrada, Mario F. Rodriguez-Moran, Cesar Arcos-Gonzalez, Jesus Mercado-Rios, Fermin Cristobal-Perez, Braulio R. Hernandez-Martinez, Marco A. Nava-Aguilar, Gilberto Gonzalez-Arroyo, Edgar P. Salazar-Fernandez, Pedro S. Quiroz-Armada, Ricarda Cortes-Vieyra, Ruth Noriega-Cisneros, Guadalupe Zinzun-Ixta, Maria C. Maldonado-Pichardo, Luis J. Flores-Alvarez, Seydhel C. Reyes-Granados, Ricardo Chagolla-Morales, Juan G. Paredes-Saralegui, Marisol Flores-Garrido, Luis M. Garcia-Velazquez, Karina M. Figueroa-Mora, Anel Gomez-Garcia, Cleto Alvarez-Aguilar, Arturo Lopez-Pineda

## Abstract

**Background:** Artificial intelligence (AI) models applied to diabetes mellitus research have grown in recent years, particularly in the field of medical imaging. However little work has been done exploring real-world data (RWD) sources such as electronic health records (EHR) mostly due to the lack of reliable public diabetes databases. However, with more than 500 million patients affected worldwide, complications of this condition have catastrophic consequences. In this manuscript we aim to first extract, clean and transform a novel diabetes research database, DiabetIA, and secondly train machine learning (ML) models to predict diabetic complications.

**Methods:** In this study, we used observational retrospective data from the Mexican Institute for Social Security (IMSS) extracting and de-identifying EHR data for almost 2 million patients seen at primary care facilities. After applying eligibility criteria for this study, we constructed a diabetes complications database. Next, we trained naïve Bayesian models with various subsets of variables, including an expert-selected model.

**Results:** The DiabetIA database is composed of 136,674 patients (414,770 records and 447 variables), with 33,314 presenting diabetes (24.3%). The most frequent diabetic complications were diabetic foot with 2,537 patients, nephropathy with 1,914 patients, retinopathy with 1,829 patients, and neuropathy with 786 patients. These complications were accurately predicted by the Gaussian naïve Bayessian models with an average area under the curve AUC of 0.86. Our expert-selected model, achieved an average AUC of 0.84 with 21 curated variables.

**Conclusion:** Our study offers the largest longitudinal research database from EHR data in Latin America for research. The DiabetIA database provides a useful resource to estimate the burden of diabetic complications on healthcare systems. Machine learning models can provide accurate estimations of the total cases presented in medical units. For patients and their clinicians, it is imperative to have a way to calculate this risk and start clinical interventions to slow down or prevent the complications of this condition.

**Brief description:** The study centers on establishing the DiabetIA database, a substantial repository encompassing de-identified electronic health records from 136,674 patients sourced from primary care facilities within the Mexican Institute for Social Security (IMSS). Our efforts involved curating, cleansing, and transforming this extensive dataset, and then employing machine learning models to predict diabetic complications with high accuracy.

## 1. Introduction

Type 2 diabetes (T2D) is a growing concern globally, affecting millions and increasing the risk of severe complications. Artificial Intelligence (AI) applications, notably supervised machine learning (ML) and deep learning (DL) models, have significantly advanced in diagnosing diabetes-related chronic complications1,2. This progress includes accurate diagnoses of conditions like diabetic retinopathy and macular edema through advanced imaging techniques, demonstrating accuracy rates close to 99%3. The need for comprehensive data sources is crucial to understand and manage diabetes effectively4. Real-world data (RWD) from surveys, claims, and particularly electronic health records (EHR), play pivotal roles in these advancements5. Real-world data (RWD) frequently presents in diverse formats, including structured, unstructured, textual, visual, or video data, each with unique challenges like high-dimensionality, missing or incomplete information, and registration errors. However, artificial intelligence has developed a variety of methods to effectively analyze such data^6^.

Diabetes mellitus, especially Type 2 Diabetes (T2D), is a prevalent metabolic disorder marked by chronic high blood sugar levels due to insufficient insulin^7^. Globally, an estimated 537 million people had diabetes in 2022, a number expected to rise to 643 million by 2030^8^. In North America, Mexico reports a significant prevalence of 18.4%, according to Mexico’s National Health and Nutrition Survey (ENSANUT)^9^. This condition poses severe complications, impacting quality of life and increasing the risk of premature death. These complications, both microvascular (nephropathy, retinopathy, neuropathy) and macrovascular (heart diseases, strokes, peripheral vascular diseases), lead to organ damage and dysfunction. Notably, diabetic nephropathy, retinopathy, and amputations due to neuropathies and arterial disease are major causes of kidney failure, blindness, and non-traumatic amputations, respectively^10,11^.

Several factors significantly impact the progression of diabetes, including being a current or former smoker, being overweight, increased waist circumference, persistent high blood pressure (even with medication), and elevated levels of key markers like HbA1c, LDL cholesterol, HDL cholesterol, triglycerides, and urinary albumin:creatinine ratio^12^. Additionally, complications such as sensory or skin changes in the feet and any form of retinopathy contribute to the disease’s complexity. As the disease evolves, patient care protocols must adapt accordingly. The evolving nature of diabetes-related hospitalizations, encompassing cases unrelated to traditional diabetes causes such as infections and cancers, underscores the necessity for retrospective studies utilizing existing databases; these analyses of disease behavior enable the implementation of innovative healthcare strategies, leading to cost reduction and improved management approaches within the healthcare system^13^.

Recent studies have yielded various algorithms for predicting chronic diabetes complications^5,14,15^. This study’s objective was to establish the DiabetIA database, drawn from a Mexican public healthcare system, constituting the largest public longitudinal research database from Electronic Health Record (EHR) data in Latin America. Unlike counterparts such as the United States’ eMERGE consortium^16^, the Million Veteran Program^17^, the MIMIC-IV database^18^, and the UK Biobank^19^, which are situated outside Latin America, DiabetIA represents a unique regional contribution. The research encompasses two primary objectives: firstly, the extraction, cleaning, and transformation of Electronic Health Record (EHR) data to construct the DiabetIA database; and secondly, the development and validation of machine learning models tailored for predicting diabetic complications. These dual goals underline the study’s commitment to creating a robust and refined dataset while leveraging advanced machine learning techniques to enhance the predictive capabilities specific to diabetic complications.

## 2. Methods

This manuscript was prepared following the TRIPOD Statement^20^ (See Supplementary Material A). Approval was obtained by the Ethics and Research Committees from the Mexican Institute of Social Security (IMSS), which are certified as Institutional Review Board (IRB). Given the secondary usage of this database the IRB waived informed consent from participants, and data was anonymized removing personal information following Mexican legislation. The protocol was registered with the number R-2018-785-051.

### 2.1 Clinical Environment and Data Source

This is a retrospective observational longitudinal cohort study in primary care units (*unidades de medicina familiar* or UMF, in Spanish) of the Mexican Institute for Social Security (IMSS), the largest healthcare institution in Mexico insuring more than half of the population. We extracted Electronic Health Records (EHR) data from six medical units, located in the state of Michoacan (western Mexico). All data presented in this study corresponds to outpatient primary care visits between 2005 and 2020. The database was deposited at the National Informatics Ecosystem repository of the National Council for Humanities, Science and Technology (CONAHCYT) and is freely accessible without registration.

### 2.2 Participants

In this study, patients from six medical units were included if they were 18 years or older and had received clinical services. For those diagnosed with Type 2 diabetes (T2D), the diagnosis had to occur after they turned 18. Inclusion required at least two consultations: one within a two-year window and another in the following year. Exclusions applied if birthdate, sex, or specific diagnoses (related to pregnancy, childbirth, or breastfeeding) were missing. Refer to Supplementary Material B for the eligibility criteria diagram. The selection of cases is described in Supplementary Material C.

For each patient, the total follow-up time was divided into three-year windows when available. Each window consisted of a two-year follow-up of data, which was used to train the predictive models, and the remaining year was used for testing. A single patient might contribute to the database one record (i.e. patients with three years of follow-up), and up to 15 records (patients with 17 years of follow-up). A detailed description of this process is provided in Supplementary Material D.

### 2.3 Variables and Missing data

In this study, predictive models employed two types of variables: predictors (X) and outcomes (Y) based on the XY model. A comprehensive list of these variables can be found in Supplementary Material E. The predictors (X) were crucially identified by medical experts, encompassing all available information from medical consultations within a two-year window. To define outcomes (Y), chronic complications of Type 2 diabetes (T2D) were pinpointed through ICD-10 codes recorded in medical consultations during the year immediately following the two-year predictor window. Specific codes like E11.2, E11.3, E11.4, and E11.5 denoted complications such as kidney issues, ophthalmic complications (most frequently retinopathy), neurological problems (commonly neuropathy), and peripheral circulatory complications (especially diabetic foot). Each complication was categorized as ’0: no complication’ if absent in both predictor and outcome windows, ’1: de novo complication’ if it emerged during the outcome window, and ’2: existing complication’ if present in both windows.

In handling missing data, diagnoses were imputed, with early diagnoses propagated for all subsequent visits even if not explicitly mentioned, ensuring continuity of medical history. Body measurements or lab values with up to 30% missing data were imputed using a multivariate iterative imputer, controlling for age, sex, diabetes, and hypertension status (all complete in the database). For other missing data, null values were assigned. Regarding medication variables, missing values were treated as missing at random and were considered not prescribed if absent in the electronic health records (EHR), contributing to a total dosage of 0 when calculating within the specified time window.

### 2.4 Computational Pipeline

In our study, we began by accessing a public data repository and performing various data engineering processes, which included removing columns without values, calculating new variables like body mass index, categorizing continuous variables using clinical knowledge and literature, and imputing missing data (details in Section 2.5). We then applied statistical methods to characterize the database at both patient and record levels, using odds ratio (OR) to determine variable importance. Additionally, we utilized t-distributed stochastic neighbor embedding (t-SNE) to visualize high-dimensional data.

For our machine learning approach, we employed a 5-fold supervised method using Gaussian naïve Bayes classifiers, a technique well-suited for Type 2 Diabetes (T2D) databases^21^. Model evaluation was performed using area under the receiver operating characteristic (AUC) metric with a 95% confidence interval and brier skilled loss (BSL) for calibration assessment. To ensure clarity, we fixed certain steps, but detailed explorations and results can be found in Supplementary Material G. Notably, we balanced classes using random undersampling, applied Yeo Johnson transformation and Z-score standardization for continuous values, and selected features based on medical knowledge. These features were categorized into demographic variables, demographic plus laboratory values, demographic plus diagnostic codes, and demographic plus prescribed medications. Statistical comparisons of AUCs between the all-variables model and other models were conducted using the DeLong method. The overall computational pipeline of our study is shown in Figure 1.

**Figure 1.**
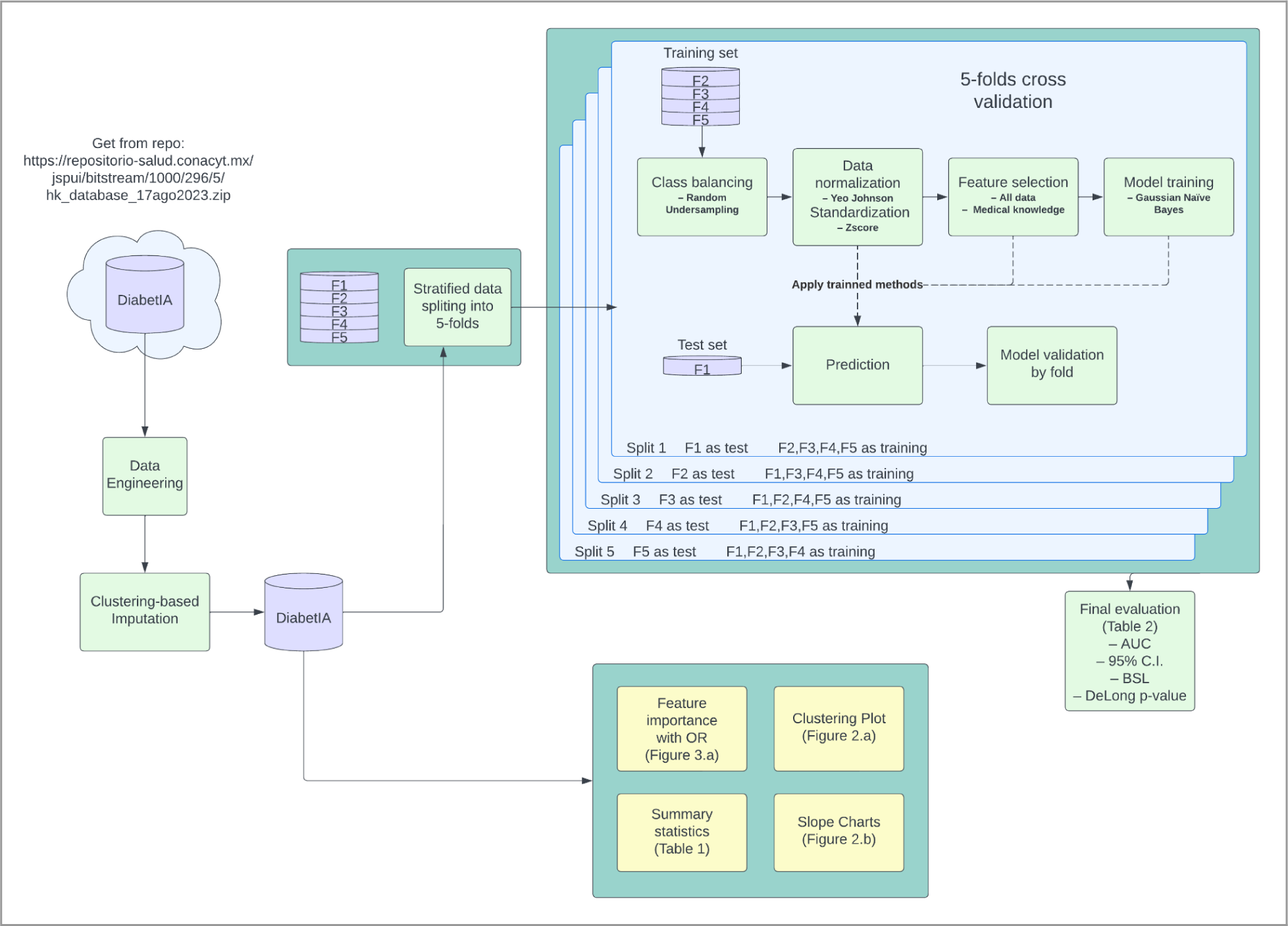
Data analysis pipeline. The DiabetIA database is extracted from a public repository. Then, data engineering and imputation is performed to build summary statistics (Table 1), feature importance of odds ratio (OR), relative risk (RR), slope charts, and clustering plot. A 5-fold machine learning process is run to train and validate the diabetic complication models.

**Table 1.** Demographic, clinical and baseline biochemical characteristics of the study population.

|  | **All patients** | **Patients without T2D** | **Patients with T2D w/o complications** | **Patients with T2D with complications** |
| --- | --- | --- | --- | --- |
|  | N (%) | N (%) | N (%) | N (%) |
| **N** | 136,674 | 103,360 | 27,297 | 6,017 |
| **Sex** |  |  |  |  |
| Female | 79,098 (57.87%) | 59,444 (57.51%) | 16,304 (59.73%) | 3,350 (55.68%) |
| Male | 57,576 (42.13%) | 43,916 (42.49%) | 10,993 (40.27%) | 2,667 (44.32%) |
| **BMI (Weight kg/Height m<sup>2</sup>)** |  |  |  |  |
| Underweight | 1,272 (0.93%) | 1,197 (1.16%) | 54 (0.20%) | 21 (0.35%) |
| Normal weight | 32,831 (24.02%) | 27,809 (26.90%) | 3,840 (14.07%) | 1,182 (19.64%) |
| Overweight | 54,400 (39.80%) | 41,106 (39.77%) | 10,808 (39.59%) | 2,486 (41.32%) |
| Obesity (any class) | 44,577 (32.62%) | 29,713 (28.75%) | 12,537 (45.93%) | 2,327 (38.67%) |
| **Age at first visit (years)** |  |  |  |  |
| 18 – 44 | 57,734 (42.24%) | 54,390 (52.62%) | 3,063 (11.22%) | 281 (4.67%) |
| 45 - 64 | 49,191 (35.99%) | 31,909 (30.87%) | 14,382 (52.69%) | 2,900 (48.20%) |
| 65 and older | 29,749 (21.77%) | 17,061 (16.51%) | 9,852 (36.09%) | 2,836 (47.13%) |
| **Age_at_dx_e11 (years)** |  |  |  |  |
| 18 – 44 | – | – | 11,575 (42.40%) | 2,724 (45.27%) |
| 45 - 64 | – | – | 12,466 (45.67%) | 2,647 (43.99%) |
| 65 and older | – | – | 3,256 (11.93%) | 646 (10.74%) |
| **Comorbidities** |  |  |  |  |
| Arterial hypertension | 50,638 (37.05%) | 25,594 (24.76%) | 20,093 (73.61%) | 4,951 (82.28%) |
| Dyslipidemia (E78) | 28,767 (21.05%) | 15,379 (14.88%) | 10,814 (39.62%) | 2,574 (42.78%) |

| **Biochemical values (mean, std)** |  |  |  |  |
| --- | --- | --- | --- | --- |
| Creatinine (mg/dL) | – | – | 0.92 ± 0.67 | 1.44 ± 1.56 |
| Glucose (mg/dL) | – | – | 137.14 ± 52.94 | 142.77 ± 48.88 |
| HbA1c (%) | – | – | 7.56 ± 1.96 | 7.80 ± 1.97 |
| Triglycerides (mg/dL) | – | – | 203.67 ± 115.75 | 199.00 ± 111.21 |
| Cholesterol (mg/dL) | – | – | 183.42 ± 40.69 | 182.29 ± 42.57 |
| **T2D Complications** |  |  |  |  |
| Nephropathy (E11.2) | – | – | – | 1,914 (31.81%) |
| Retinopathy (E11.3) | – | – | – | 1,829 (30.40%) |
| Neuropathy (E11.4) | – | – | – | 786 (13.06%) |
| Diabetic foot (E11.5) | – | – | – | 2,537 (42.16%) |

T2D= Type 2 diabetes; BMI= Body Mass Index; Underweight= BMI<18; Normal weight= BMI between 18 to 24.9; Overweight= BMI ≥25 and <30; Obesity = BMI ≥30. Arterial hypertension = Systolic blood pressure values ≥140 mm/Hg and diastolic blood pressure ≥90 mm/Hg, or lower values under treatment with antihypertensive drugs. Dyslipidemia= Serum concentrations of total cholesterol >200 mg/dL and/or Triglycerides >150 mg/dL; or HDL cholesterol <40 mg/dL in men and <50 mg/dL in women. HbA1c= Hemoglobin-A1c.

## 3. Results

The DiabetIA database is composed of 136,674 patients which are summarized in Table 1. This database was compiled from visits in IMSS primary care units in Michoacan, a western Mexican state with 4.7 million inhabitants by 2020^22^. Given that IMSS insures and provides healthcare to half of the Mexican population, our study can be considered the closest to a state-wide census. Table 1 shows the demographic, clinical and biochemical characteristics of the study population, stratifying by their status of diabetes without and with complications. It is observed that the frequency of T2D was more common in the population aged 45 years and older (57.60%); however, the high frequency found (42.40%) in the younger population is also striking, in patients with and without complications. The same trend is observed in the age of the population with T2D with the presence of chronic complications, with almost equal proportions of women and men, with an absolute predominance of overweight and obesity, since only 26.9% of the population without diabetes and 33.71% of the population with diabetes has a registered normal weight. Arterial hypertension and dyslipidemia are the main documented comorbidities, and a chronic metabolic imbalance in patients with T2D characterized by high serum concentrations of fasting glucose, high percentage (%) of HbA_1c_, as well as high levels of triglycerides. The chronic complications recorded were predominantly diabetic foot (42.16%) followed by diabetic nephropathy (31.81%) and diabetic retinopathy (30.40%).

We investigated further the clinical characteristics of patients with T2D complications and summarized them in Figure 2 (A-D). Slope plots are shown for the main laboratory values, which reflects differences among the *de novo* and existing complications groups. However, in most cases the difference is marginal, and cannot be ascertained only by observation of the plots. These small differences are better suited to be interpreted by machine learning models. Patient similarity plots are shown in the form of TSNe plots, differentiating by complication. The patterns shown in Figure 2 (E-H) reflect the subclusters that exist within this cohort. All the existing and de novo patients are grouped together, regardless of whether they are de novo or pre-existing complication patients (shown in color). Meanwhile, those patients without complications are grouped in separate clusters (shown in gray).

**Figure 2.**
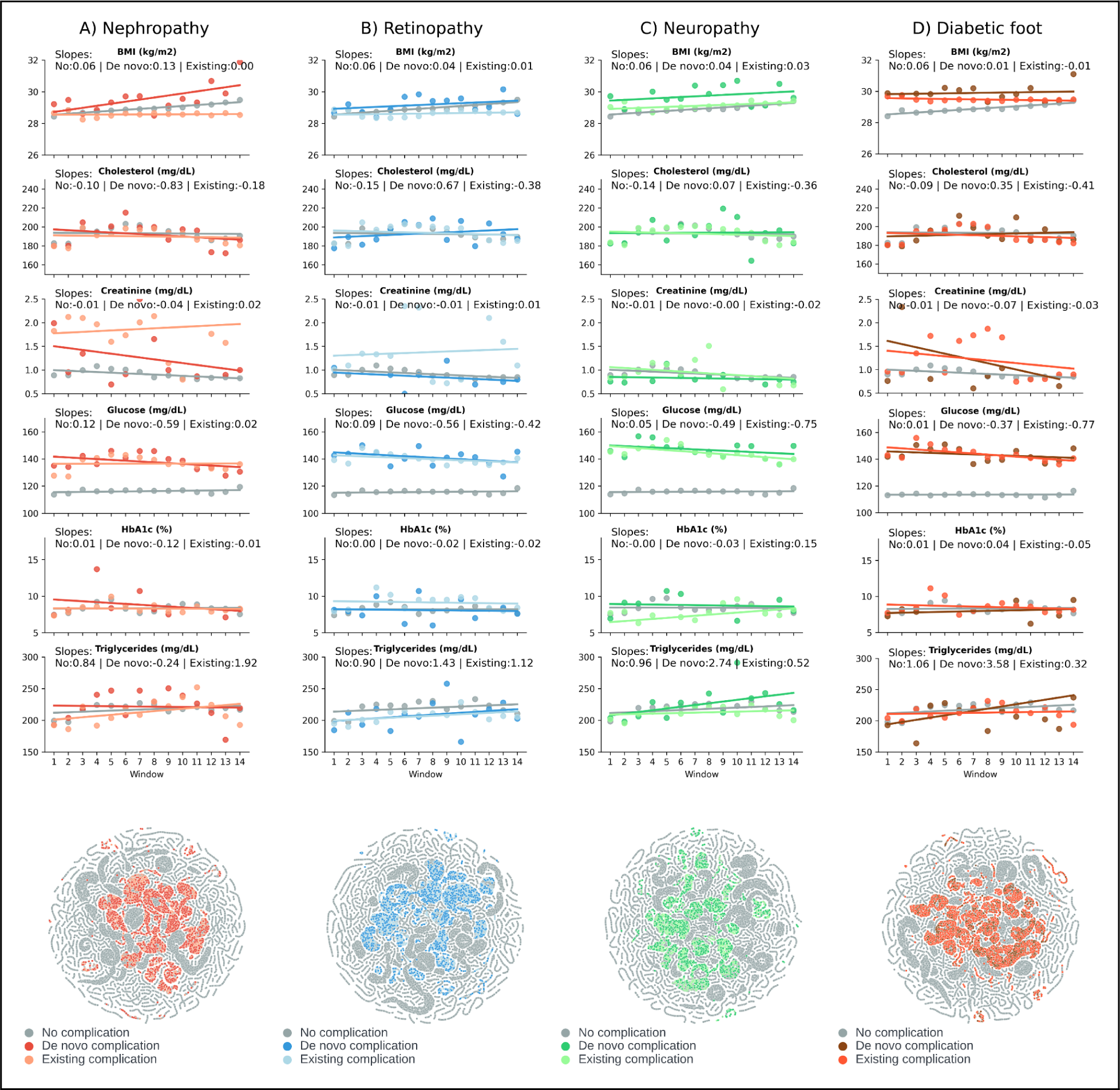
Slope and TSNe plots for the main T2D complications (in columns). The top row shows slope plots for BMI, total cholesterol, creatinine, glucose, HbA1c, and triglycerides. The bottom row shows the TSNe plots for each T2D complication.

The odds ratio (OR) measures the probability of an event happening in one group compared to another, enabling a comparison between participants with de novo complications and those with existing complications. This comparison is illustrated in Figure 3 (a-d) using the variables selected in the expert-selected model, while the graphical representation of this model is depicted in Figure 3(e). The predictive results, measured as AUC, for all naïve Bayesian models and its associated statistics are shown in Table 2. In addition to naïve Bayesian models, we also built other machine learning models, which we presented in Supplementary Material G.

**Figure 3.**
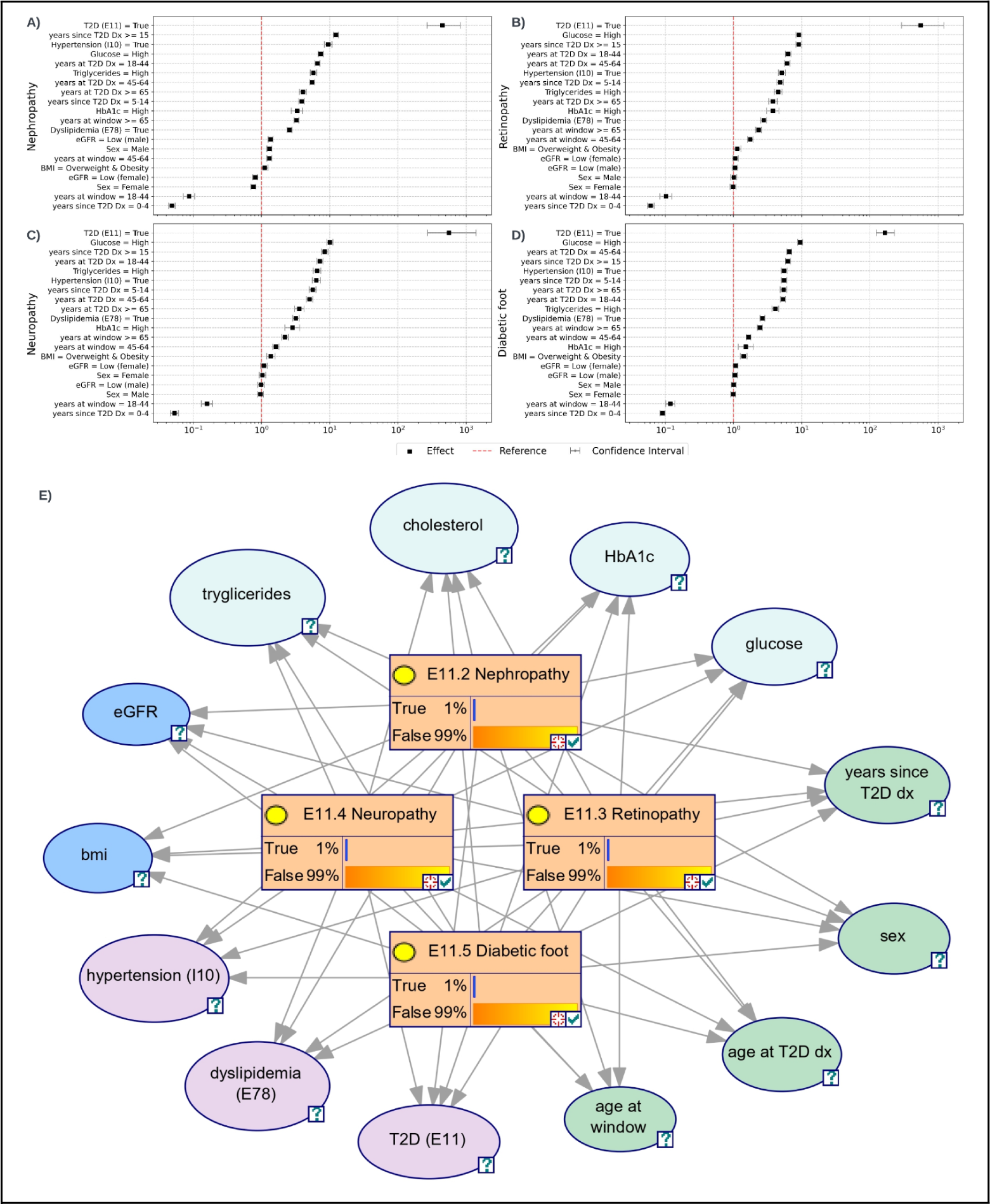
Sections A-D display the odds ratio (OR) forest plot comparing participants with de novo complications to those with existing complications. In Section E, the expert-selected naïve Bayesian model is visually depicted: green nodes denote demographic variables, pink nodes represent current diagnoses, light blue indicate laboratory values, dark blue indicates calculated variables from other metrics, and orange nodes signify target chance nodes, reflecting a priori probabilities for each diabetic complication (less than 1%). As evidence accrues for each patient, these probabilities are updated using Bayes’ theorem, facilitating dynamic visualization of the model’s predictive accuracy. Acronyms: T2D: type 2 diabetes, HTN: arterial hypertension, BMI: body mass index, HbA1c: Hemoglobin A1c, Dx: diagnosis, Wx: 2-year time window.

**Table 2.** Machine learning results for each T2D complication. Model 1, utilizes all 447 variables in the database and serves as the benchmark. Model 2 incorporates 13 variables expert-selected by physicians but with machine-learned weights. Model 3 was based solely on demographic variables. Models 4, 5, and 6, integrate demographic variables expanded with laboratory values, diagnoses, or drugs respectively. DeLong’s method was used for estimating confidence intervals.

|  | **Nephropathy** | **Retinopathy** | **Neuropathy** | **Diabetic Foot** |
| --- | --- | --- | --- | --- |
| **Model 1: All variables model**<br>**(vars = 447)** |  |  |  |  |
| AUC | 0.86 | 0.86 | 0.86 | 0.85 |
| 95% C.I. | (0.85, 0.86) | (0.85, 0.86) | (0.85, 0.86) | (0.85, 0.85) |
| BSL | 0.23 | 0.23 | 0.24 | 0.24 |
| **Model 2: Expert-selected model**<br>**(vars = 13)** |  |  |  |  |
| AUC | 0.84 | 0.84 | 0.83 | 0.84 |
| 95% C.I. | (0.83, 0.84) | (0.84, 0.85) | (0.82, 0.83) | (0.83, 0.84) |
| BSL | 0.18 | 0.19 | 0.18 | 0.18 |
| P-value relative to Model 1 | < 0.001 | <0.001 | <0.001 | <0.001 |
| **Model 3: Demographic-only model**<br>**(vars = 9)** |  |  |  |  |
| AUC | 0.83 | 0.82 | 0.81 | 0.82 |
| 95% C.I. | (0.82, 0.83) | (0.81, 0.82) | (0.800, 0.81) | (0.82, 0.83) |
| BSL | 0.18 | 0.19 | 0.19 | 0.20 |
| P-value relative to Model 1 | < 0.001 | <0.001 | <0.001 | <0.001 |
| **Model 4: Demographic + Labs model**<br>**(vars = 115)** |  |  |  |  |
| AUC | 0.82 | 0.87 | 0.84 | 0.87 |
| 95% C.I. | (0.81, 0.82) | (0.86, 0.87) | (0.83, 0.84) | (0.86, 0.87) |
| BSL | 0.16 | 0.24 | 0.19 | 0.23 |
| P-value relative to c | < 0.001 | <0.001 | <0.001 | <0.001 |
| **Model 5: Demographic + Diagnoses**<br>**model**<br>**(vars = 163)** |  |  |  |  |
| AUC | 0.86 | 0.87 | 0.87 | 0.86 |
| 95% C.I. | (0.86, 0.87) | (0.86, 0.87) | (0.86, 0.87) | (0.86, 0.86) |
| BSL | 0.25 | 0.24 | 0.25 | 0.24 |
| P-value relative to Model 1 | < 0.001 | 0.002 | 0.002 | 0.042 |
| **Model 6: Demographic + Drugs model**<br>**(vars = 119)** |  |  |  |  |
| AUC | 0.86 | 0.86 | 0.86 | 0.84 |
| 95% C.I. | (0.85,0.86) | (0.85, 0.86) | (0.85, 0.86) | (0.83, 0.84) |
| BSL | 0.23 | 0.24 | 0.24 | 0.19 |
| P-value relative to Model 1 | 0.644 | 0.806 | 0.604 | <0.001 |

## 4. Discussion

### The DiabetIA database and the machine learning models

In this study, we introduce the DiabetIA database, a comprehensive longitudinal dataset sourced from EHRs at primary care facilities under the IMSS healthcare system in Michoacan, Mexico. IMSS, the country’s largest healthcare provider, covers half the population through a blend of employee, employer, and government funding sources. DiabetIA stands as a pivotal research resource, openly accessible in a public repository, dedicated to advancing diabetes knowledge and technology through artificial intelligence (AI). Our research utilized naïve Bayesian models to successfully predict diabetic complications up to one year in advance. Notably, our models achieved exceptional accuracy, with an AUC surpassing 0.82 across all complications (peaking at 0.87), a performance level superior to any reported in existing literature for this task. We explored diverse subsets of variables, including demographics, laboratory data, prescription information, and ICD-10 diagnoses, revealing nuanced insights into their predictive potential. This pioneering work not only expands our understanding of diabetes but also offers a robust framework for future AI-driven advancements in the field.

### Clinical implications of ML predictive models of diabetic complications

Our expert-selected model offers a practical solution, striking a balance between predictive accuracy and the number of variables used. Its applicability in real-world settings, particularly within primary care units, is evident, providing healthcare practitioners with an innovative and efficient tool.

Type 2 diabetes ranks among the most prevalent chronic diseases, second only to hypertension. It stands as a primary cause of chronic kidney disease (CKD) and end-stage CKD, with HTN being a common comorbidity^24^. Notably, diabetic retinopathy, a leading cause of vision loss globally, affects individuals between 25 and 74 years with T2D. Diabetic neuropathy, primarily afflicting the lower extremities, manifests in various degrees of severity, from mild discomfort to debilitating pain and life-threatening complications. Symptoms include pain, leg numbness, and disruptions in digestion, bladder, and heart rate control^25^. Moreover, diabetic foot complications are a significant source of morbidity, contributing to two-thirds of non-traumatic amputations in the United States^26^. According to the World Health Organization, diabetes-related complications in the lower extremities rank among the top ten conditions in terms of years lived with disability^27^.

T2D results from a complex interplay of non-modifiable factors like heredity and age, along with modifiable elements such as overweight, obesity, physical inactivity, high blood pressure, and dyslipidemia, all contributing to the risk of chronic complications associated with T2D^28^. While early detection and cutting-edge treatments have notably enhanced disease management and patients’ quality of life, there’s an imperative to explore innovative methods leveraging technology like AI. These approaches can empower multidisciplinary healthcare teams and individuals, enabling more effective management of health. AI offers pioneering solutions in prevention, diagnosis, and treatment, facilitating swift and scalable automation, extending far beyond mere glycemic control.

### Impact on public health in Mexico and the World

Recent data from the World Health Organization (WHO) underscores the alarming rise of T2D in economically disadvantaged nations, notably in countries like Mexico. Our analysis of the DiabetIA database mirrors these trends, revealing a concerning increase in T2D among younger demographics, a departure from its historically prevalent occurrence in middle-aged and older individuals. This shift in disease demographics, as indicated in our study and corroborated by global analyses^31^ presents a pivotal challenge.

The global rise in T2D poses a substantial challenge for healthcare systems worldwide. If the current exponential increase in T2D cases persists, no healthcare system will be able to bear the economic burden associated with T2D and its complications. Therefore, it is crucial to have advanced technological tools like the one developed by our group. These tools enable the identification of individuals at high risk and facilitate the early diagnosis of chronic diabetes complications. Implementing effective interventions using such technology is essential to alter the natural course of diabetes and its associated complications.

### Study Limitations and Strengths

A potential drawback of our study lies in the absence of external validation, which is essential for assessing its adaptability across diverse clinical scenarios and different patient demographics. Additionally, our algorithm, while promising, hasn’t been validated in clinical practice, raising questions about its real-world efficacy and reliability. Addressing these challenges is paramount in ensuring the algorithm’s robustness and practicality.

Conversely, our expert-selected model boasts notable strengths, including the careful selection of variables, which are both parsimonious and explainable. This deliberate selection not only aligns with the current understanding of diabetes complications but also ensures the algorithm’s future applicability and ease of interpretation in complex healthcare environments.

### Conclusion

Our study demonstrates the transformative potential of leveraging RWD to deepen our understanding of diabetes. Through the application of machine learning models, we have achieved remarkable success in identifying de novo diabetic complications, showcasing the power of Artificial Intelligence in diabetes research. This achievement not only fulfills the promise of AI but also augments the capabilities of human scientists, marking a significant advancement in the field. Our expert-selected model developed in this study exhibits promising features that make it suitable for deployment in local healthcare settings. Its ability to accurately predict diabetic complications underscores its clinical relevance and potential to enhance patient outcomes.

Finally, our study’s pivotal contribution lies in the creation of the DiabetIA database. The meticulous processes of extraction, transformation, and publication have resulted in a comprehensive and valuable resource. We expect the DiabetIA database to become an indispensable resource for researchers globally, fostering deeper explorations and breakthroughs at the intersection of AI, healthcare, and diabetes research, thereby promising a transformative future for medical science.

## Acknowledgements

Competing interests

Authors ALP and PSQA hold shares of Amphora Health. Authors JT, DSQ, MFRM, MANA, EPSF, GGA contributed to the research while employed by Amphora Health. Authors RCM, JGPS, AGG are employed by IMSS. All other authors from academic institutions declare no conflict of interest.

## Contributions

ALP and CAA designed the study. JTG, DSQ, RPE, performed computational cleaning and transformation of data. JT, DSQ, RPE, CAG, JMR, FCP, BRHM, MANA, GGA, EPSF, PSQA provided computational programming and machine learning support. RCV, RNC, GZI, MCMP, LJFA provided clinical curation of data. MFRM, CAA selected the variables of the expert-selected model. RCM, JGPS, AGG, extracted the dataset from the EHR and anonymized the dataset. JT, MFRM, SCRG, MFG, LMGV, KMFM, AGG, CAA, ALP provided interpretation of the results. ALP and CAA drafted the manuscript, a large language model (LLM) was used to assist in copy-editing this manuscript, and all human co-authors contributed critically, read, revised and approved the final version.

## Ethics approval

This study was approved by the Ethics and Research Committees from the Mexican Institute of Social Security (IMSS), which are certified as Institutional Review Board (IRB) in accordance with the Mexican regulation, under protocol numbers R-2018-785-051. Since, this was a retrospective study and the author AGG and RCM carried out the anonymization of data, the IRB waived the need for consent for this study.

## Data Availability

The data that supports the findings of this study is available for research purposes at the National Informatics Ecosystem under its Health chapter (ENI Salud), which can be freely accessed at: https://repositorio-salud.conacyt.mx/jspui/handle/1000/56

## Code Availability

The programming code used in this study is available in the Amphora Health public repository: https://github.com/AmphoraHealth/diabetia

