## Supplementary Material for "DiabetIA: Building Machine Learning Models for Type 2 Diabetes Complications"

### DiabetIA: a real-world research database to predict de novo diabetic complications using artificial intelligence.

#### Supplementary Material

#### A. TRIPOD Statement

| Section/Topic | Item | Checklist Item | Page |
|---|---|---|---|
| **Title and abstract** | | | |
| **Title** | 1 | Identify the study as developing and/or validating a multivariable prediction model, the target population, and the outcome to be predicted. | Title mentions machine learning, diabetes complications, and patients. |
| **Abstract** | 2 | Provide a summary of objectives, study design, setting, participants, sample size, predictors, outcome, statistical analysis, results, and conclusions. | All components mentioned. |
| **Introduction** | | | |
| **Background and objectives** | 3a | Explain the medical context (including whether diagnostic or prognostic) and rationale for developing or validating the multivariable prediction model, including references to existing models. | Introduction Section, paragraph 1, 2. |

| | | | |
|---|---|---|---|
| | 3b | Specify the objectives, including whether the study describes the development or validation of the model or both. | Introduction Section, paragraph 4 |
| **Methods** | | | |
| **Source of data** | 4a | Describe the study design or source of data (e.g., randomized trial, cohort, or registry data), separately for the development and validation data sets, if applicable. | Methods Section, subsection 2.1 Clinical Environment and Data Source |
| | 4b | Specify the key study dates, including start of accrual; end of accrual; and, if applicable, end of follow-up. | Methods Section, subsection 2.1 Clinical Environment and Data Source |
| **Participants** | 5a | Specify key elements of the study setting (e.g., primary care, secondary care, general population) including number and location of centers. | Methods Section, subsection 2.2 Participants |
| | 5b | Describe eligibility criteria for participants. | Methods Section, subsection 2.3 Participants |
| | 5c | Give details of treatments received, if relevant. | NA |
| **Outcome** | 6a | Clearly define the outcome that is predicted by the prediction model, including how and when assessed. | Methods Section, subsection 2.4 Computational Pipeline |
| | 6b | Report any actions to blind assessment of the outcome to be predicted. | Methods Section, subsection 2.4 Computational Pipeline |
| **Predictors** | 7a | Clearly define all predictors used in developing or validating the multivariable prediction model, including how and when they were measured. | Methods Section, subsection 2.4 Computational Pipeline |
| | 7b | Report any actions to blind assessment of predictors for the outcome and other predictors. | Methods Section, subsection 2.4 Computational Pipeline |
| **Sample size** | 8 | Explain how the study size was arrived at. | Methods Section, subsection 2.3 Participants |
| **Missing data** | 9 | Describe how missing data were handled (e.g., complete-case analysis, single imputation, multiple imputation) with details of any imputation method. | Methods Section, subsection 2.3 Variables and Missing data |

| | | | |
|---|---|---|---|
| **Statistical analysis methods** | 10a | Describe how predictors were handled in the analyses. | Methods Section, subsection 2.3 Variables and Missing data |
| | 10b | Specify type of model, all model-building procedures (including any predictor selection), and method for internal validation. | Methods Section, subsection 2.3 Variables and Missing data |
| | 10d | Specify all measures used to assess model performance and, if relevant, to compare multiple models. | Methods Section, subsection 2.3 Variables and Missing data |
| **Risk groups** | 11 | Provide details on how risk groups were created, if done. | NA |
| **Results** | | | |
| **Participants** | 13a | Describe the flow of participants through the study, including the number of participants with and without the outcome and, if applicable, a summary of the follow-up time. A diagram may be helpful. | Results Section, paragraph 1 |
| | 13b | Describe the characteristics of the participants (basic demographics, clinical features, available predictors), including the number of participants with missing data for predictors and outcome. | Results Section, Table 1 |
| **Model development** | 14a | Specify the number of participants and outcome events in each analysis. | Results Section, Table 1 |
| | 14b | If done, report the unadjusted association between each candidate predictor and outcome. | NA |
| **Model specification** | 15a | Present the full prediction model to allow predictions for individuals (i.e., all regression coefficients, and model intercept or baseline survival at a given time point). | Results Section, Figure 3 |
| | 15b | Explain how to use the prediction model. | Results Section, Figure 3 footer |
| **Model performance** | 16 | Report performance measures (with CIs) for the prediction model. | Results Section, Table 2 |
| **Discussion** | | | |
| **Limitations** | 18 | Discuss any limitations of the study (such as non representative sample, few events per predictor, missing data). | Discussion Section, subsection: Study Limitations and Strengths |

| | | | |
|---|---|---|---|
| **Interpretation** | 19b | Give an overall interpretation of the results, considering objectives, limitations, and results from similar studies, and other relevant evidence. | Discussion Section, subsection: The DiabetIA database and the machine learning models |
| **Implications** | 20 | Discuss the potential clinical use of the model and implications for future research. | Discussion Section, subsection: Clinical implications of ML predictive models of diabetic complications |
| **Other information** | | | |
| **Supplementary information** | 21 | Provide information about the availability of supplementary resources, such as study protocol, Web calculator, and data sets. | Acknowledgements Section, subsections: Data Availability and Code Availability |
| **Funding** | 22 | Give the source of funding and the role of the funders for the present study. | Acknowledgements Section, subsections: Funding |

### B. Eligibility Criteria Diagram

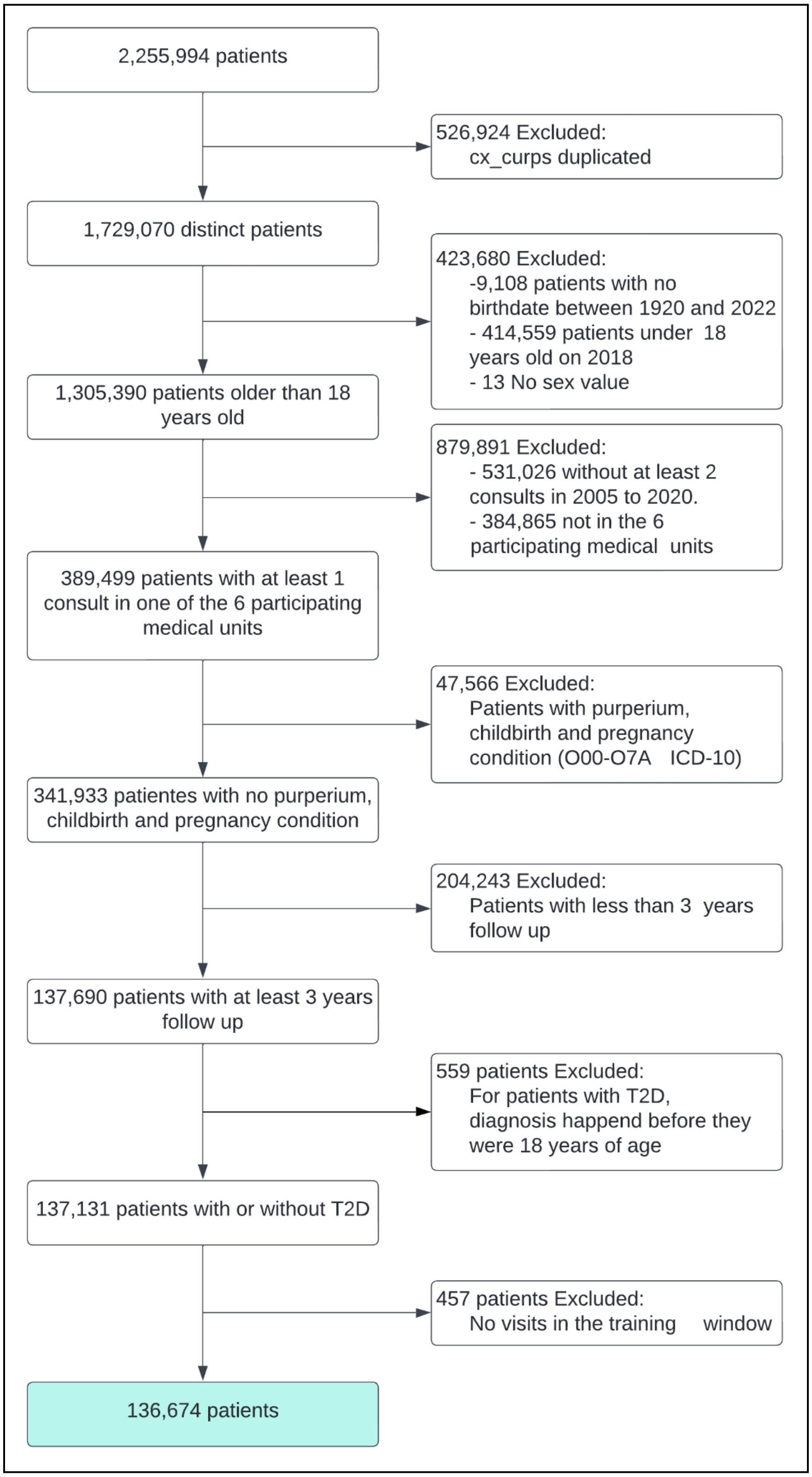

#### C. Case selection algorithm

A patient was considered to have diabetes if they had a recorded ICD-10 diagnosis of code E11 or any derived coding (e.g. E11.9). In addition, IMSS maintains an institutional census of patients with diabetes, which was useful to estimate those patients whose diagnosis of diabetes pre-dates the beginning of the EHR implementation in 2004. Patients in this census were included in the T2D group, even if they were not registered with an E11 diagnosis. The diagnosis year was selected first from the institutional census and then from the first E11 recorded.

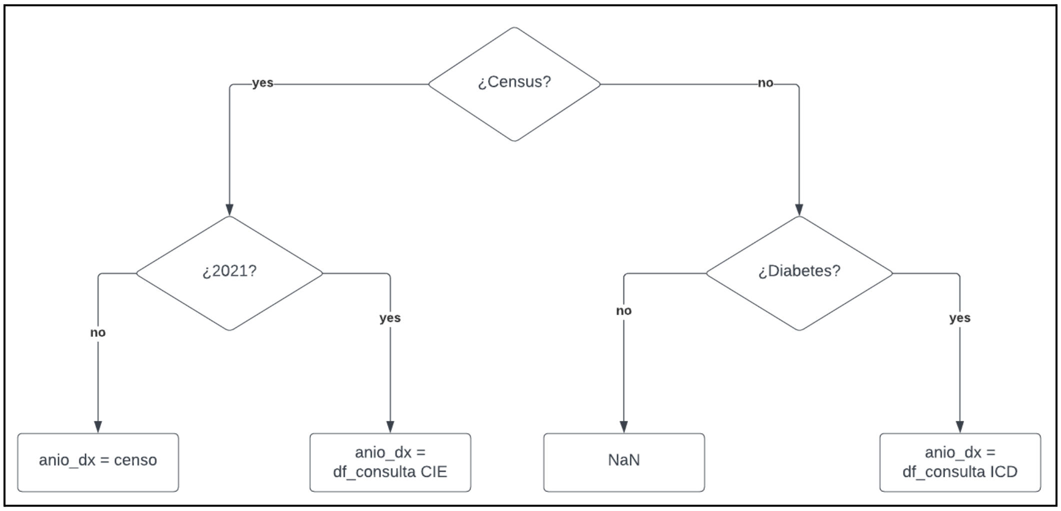

Algorithm to choose first patient diabetes diagnosis date

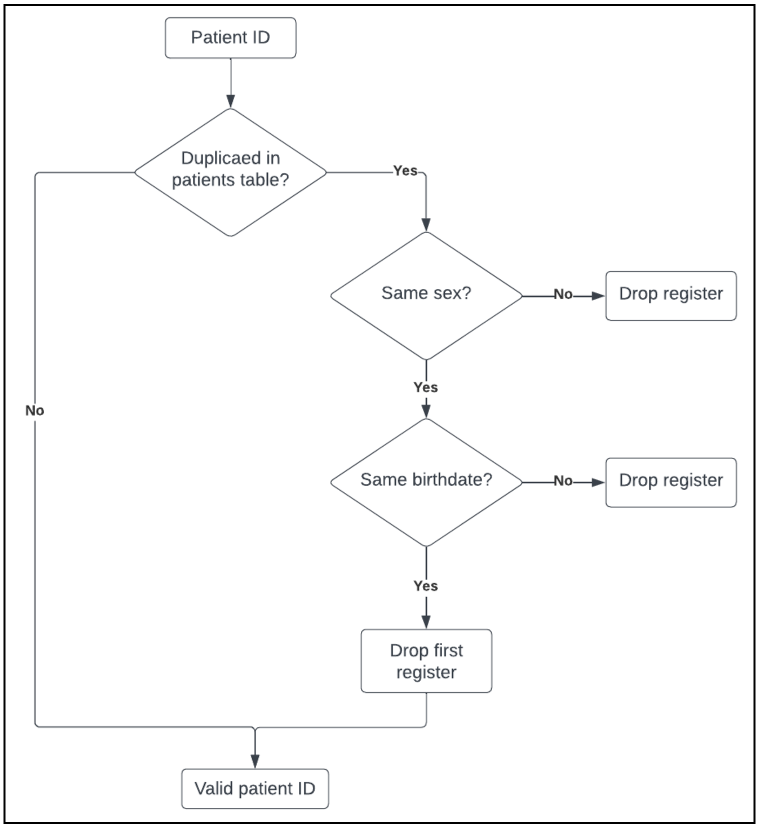

Algorithm to identify and drop duplicated patients ID

### D. Preprocessing of raw data into three-year window records

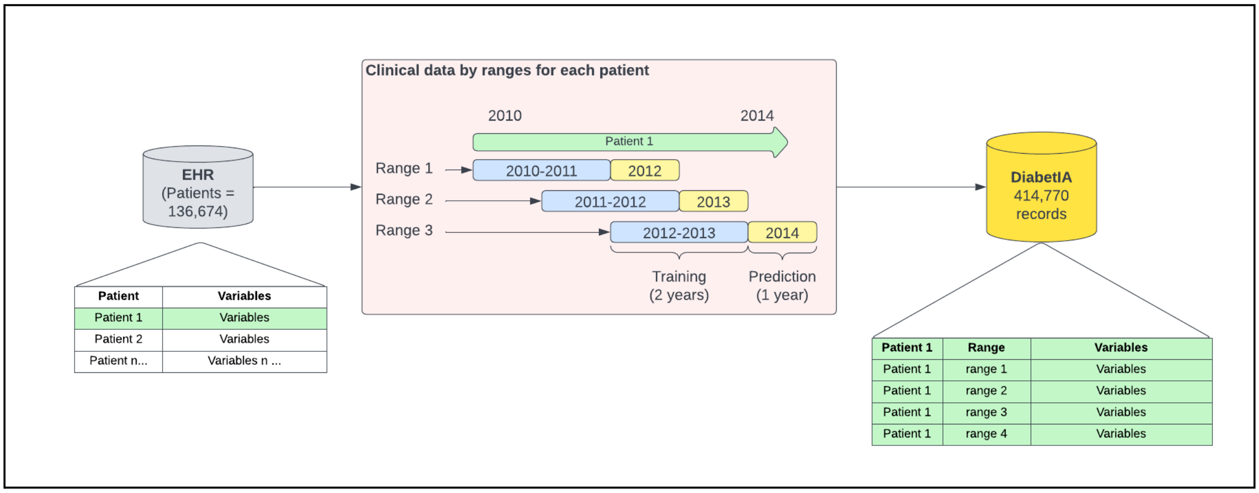

### E. List of Variables

Diagnosis variables. These were recorded at the ICD-10 chapter level (e.g. neoplasms for all C00 to D49 codes). The circulatory and endocrine system chapters are recorded at the third level of the ICD-10 hierarchy (e.g. E10 type 1 diabetes mellitus). Finally, the diabetes groups were recorded as variables in the fourth level of the ICD-10 hierarchy (e.g. E11.2, E11.3, E11.4, E11.5).

| Group | Variable | Type | Description |
|---|---|---|---|
| Demographics (17) | id | String | Unique ID for patient by window |
| | birthdate | Integer | Ordinal birthdate of patient |
| | years_since_dx | Float | Years since type 2 diabetes mellitus 2 was diagnosticated |
| | cs_sex | Integer | Sex of patient (0=Female and 1=Male) |
| | hgz_con_mf_no_12 | Binary | Family Medicine Unit located in Lázaro Cárdenas, Michoacán, México |
| | hgz_con_mf_no_2 | Binary | Family Medicine Unit loacted in Zacapu, Michoacán, México |
| | umf_no_75 | Binary | Family Medicine Unit loacted in Morelia, Michoacán, México |
| | umf_no_76 | Binary | Family Medicine Unit loacted in Uruapan, Michoacán, México |
| | umf_no_77 | Binary | Family Medicine Unit loacted in La Piedad, Michoacán, México |
| | umf_no_80 | Binary | Family Medicine Unit loacted in Morelia, Michoacán, México |
| | count_cx_w | Float | Number of visits |
| | age_at_wx | Integer | Age at the start of the window |
| | age_at_wx_label | String | Group of age at the start of the window |
| | age_at_wx_ordinal | Integer | Ordinal group of age at the start of the window |
| | dx_age_e11 | Float | Age at first type 2 diabetes mellitus diagnosis |
| | dx_age_e11_label | String | Age group at first type 2 diabetes meliitus diagnosis |
| | dx_age_e11_ordinal | Integer | Ordinal age group at first type 2 diabetes mellitus diagnosis |
| Medical diagnosis (160) | diabetes_mellitus_type_2 | Binary | Type 2 diabetes mellitus (ICD E11) |
| | essential_(primary)_hypertension | Binary | Essential (primary) hypertension (ICD I10) |

| | | | |
|---|---|---|---|
| | circulatory_system_diseases | Binary | Circulatory system diseases (ICD I00) |
| | endocrine_nutritional_and_metabolic_diseases | Binary | Nutritional and metabolic endocrine diseases (ICD E00) |
| | diseases_of_the_musculoskeletal_system_and_connective_tissue | Binary | Diseases of the musculoskeletal system and connective tissue (ICD M00) |
| | nervous_system_diseases | Binary | Nervous system diseases (ICD G00) |
| | factors_influencing_health_status_and_contact_with_health_services | Binary | Factors influencing health status and contact with health services (ICD Z00) |
| | diseases_of_the_genitourinary_system | Binary | Diseases of the genitourinary system (ICD N00) |
| | diseases_of_the_respiratory_system | Binary | Diseases of the respiratory system (ICD J00) |
| | diseases_of_the_eye_and_its_annexes | Binary | Diseases of the eye and its annexes (ICD H00) |
| | mental_and_behavioral_disorders | Binary | Mental and behavioral disorders (ICD F00) |
| | digestive_system_diseases | Binary | Digestive system diseases (ICD K00) |
| | symptoms_signs_and_abnormal_clinical_and_laboratory_findings_not_elsewhere_classified | Binary | Symptoms signs and abnormal clinical and laboratory findings not elsewhere classified (ICD R00) |
| | ear_and_mastoid_disease | Binary | Ear and mastoid disease (ICD H60) |
| | certain_infectious_and_parasitic_diseases | Binary | Certain infectious and parasitic diseases (ICD A00) |
| | skin_and_subcutaneous_tissue_diseases | Binary | Skin and subcutaneous tissue diseases (ICD L00) |
| | injuries_poisoning_and_some_other_consequences_of_external_causes | Binary | Injuries, poisoning and some other consequences of external causes (ICD S00) |
| | tumors_(neoplasms) | Binary | Tumors (neoplasms) (ICD C00) |
| | congenital_malformations_deformities_and_chromosomal_abnormalities | Binary | Congenital malformations, deformities and chromosomal abnormalities (ICD Q00) |
| | diseases_of_the_blood_and_blood_forming_organs_and_certain_disorders_affecting_the_immune_mechanism | Binary | Diseases of the blood and blood-forming organs and certain disorders affecting the immune mechanism (ICD D50) |
| | causes_for_special_purposes | Binary | Causes for special purposes (ICD U04) |
| | external_causes_of_morbidity_and_mortality | Binary | External causes of morbidity and mortality (ICD V01) |
| | certain_conditions_originating_in_the_perinatal_period | Binary | Certain conditions originating in the perinatal period (ICD P00) |
| | hypotension | Binary | hypotension (ICD I95) |

| | | | |
|---|---|---|---|
| | other_cardiac_arrhythmias | Binary | Other cardiac arrhythmias (ICD I49) |
| | hypertensive_heart_disease | Binary | hypertensive heart disease (ICD I11) |
| | hypertensive_kidney_disease | Binary | hypertensive kidney disease (ICD I12) |
| | complications_and_ill_defined_descriptions_of_heart_disease | Binary | Complications and ill-defined descriptions of heart disease (ICD I51) |
| | other_vein_disorders | Binary | Other vein disorders (ICD I87) |
| | sequelae_of_cerebrovascular_disease | Binary | Sequelae of cerebrovascular disease (ICD I69) |
| | hemorrhoids | Binary | Hemorrhoids (ICD I84) |
| | nonspecific_lymphadenitis | Binary | Nonspecific lymphadenitis (ICD I88) |
| | other_peripheral_vascular_diseases | Binary | Other peripheral vascular diseases (ICD I73) |
| | varicose_veins_of_the_lower_limbs | Binary | Varicose veins of the lower limbs (ICD I83) |
| | endocarditis_unspecified_valve | Binary | Endocarditis unspecified valve (ICD I38) |
| | chronic_ischemic_heart_disease | Binary | Chronic ischemic heart disease (ICD I25) |
| | other_conduction_disorders | Binary | Other conduction disorders (ICD I45) |
| | acute_cerebrovascular_accident_not_specified_as_hemorrhagic_or_ischemic | Binary | Acute cerebrovascular accident not specified as hemorrhagic or ischemic (ICD I64) |
| | other_and_unspecified_disorders_of_the_circulatory_system | Binary | Other and unspecified disorders of the circulatory system (ICD I99) |
| | diseases_of_the_capillaries | Binary | Diseases of the capillaries (ICD I78) |
| | acute_myocardial_infarction | Binary | acute myocardial infarction (ICD I21) |
| | aortic_aneurysm_and_dissection | Binary | Aortic aneurysm and dissection (ICD I71) |
| | phlebitis_and_thrombophlebitis | Binary | Phlebitis and thrombophlebitis (ICD I80) |
| | varicose_veins_from_other_sites | Binary | Varicose veins from other sites (ICD I86) |
| | paroxysmal_tachycardia | Binary | Paroxysmal tachycardia (ICD I47) |
| | atrial_fibrillation_and_flutter | Binary | Atrial fibrillation and flutter (ICD I48) |
| | pulmonary_embolism | Binary | Pulmonary embolism (ICD I26) |
| | other_aneurysms_and_dissecting_aneurysms | Binary | Other aneurysms and dissecting aneurysms (ICD I72) |
| | esophageal_varices | Binary | Esophageal varices (ICD I85) |
| | multiple_valve_diseases | Binary | Multiple valve diseases (ICD I08) |

| | | | |
|---|---|---|---|
| | rheumatic_diseases_of_the_mitral_valve | Binary | Rheumatic diseases of the mitral valve (ICD I05) |
| | heart_failure | Binary | Heart failure (ICD I50) |
| | cardiomyopathy | Binary | Cardiomyopathy (ICD I42) |
| | nonrheumatic_mitral_valve_disorders | Binary | Nonrheumatic mitral valve disorders (ICD I34) |
| | other_embolisms_and_venous_thrombosis | Binary | Other embolisms and venous thrombosis (ICD I82) |
| | other_cardiopulmonary_diseases | Binary | Other cardiopulmonary diseases (ICD I27) |
| | cerebral_stroke | Binary | Cerebral stroke (ICD I63) |
| | occlusion_and_stenosis_of_the_cerebral_arteries_without_causing_cerebral_infarction | Binary | Occlusion and stenosis of the cerebral arteries without causing cerebral infarction (ICD I66) |
| | atrioventricular_and_left_bundle_branch_block | Binary | Atrioventricular and left bundle branch block (ICD I44) |
| | other_cerebrovascular_diseases | Binary | Other cerebrovascular diseases (ICD I67) |
| | angina_pectoris | Binary | Angina pectoris (ICD I20) |
| | non_rheumatic_disorders_of_the_aortic_valve | Binary | Nonrheumatic disorders of the aortic valve (ICD I35) |
| | occlusion_and_stenosis_of_the_precerebral_arteries_without_causing_cerebral_infarction | Binary | Occlusion and stenosis of the precerebral arteries without causing cerebral infarction (ICD I65) |
| | rheumatic_diseases_of_the_tricuspid_valve | Binary | Rheumatic diseases of the tricuspid valve (ICD I07) |
| | arterial_embolism_and_thrombosis | Binary | Arterial embolism and thrombosis (ICD I74) |
| | other_arterial_or_arteriolar_disorders | Binary | Other arterial or arteriolar disorders (ICD I77) |
| | secondary_hypertension | Binary | Secondary hypertension (ICD I15) |
| | other_diseases_of_the_pulmonary_vessels | Binary | Other diseases of the pulmonary vessels (ICD I28) |
| | other_rheumatic_heart_diseases | Binary | Other rheumatic heart diseases (ICD I09) |
| | rheumatic_diseases_of_the_aortic_valve | Binary | Rheumatic diseases of the aortic valve (ICD I06) |
| | atherosclerosis | Binary | Atherosclerosis (ICD I70) |
| | rheumatic_fever_with_cardiac_involvement | Binary | Rheumatic fever with cardiac complication (ICD I01) |
| | other_non_infectious_disorders_of_the_lymph_vessels_and_lymph_nodes | Binary | Other non-infectious disorders of lymph vessels and lymph nodes (ICD I89) |

| | | | |
|---|---|---|---|
| | other_diseases_of_the_pericardium | Binary | Other diseases of the pericardium (ICD I31) |
| | intraencephalic_hemorrhage | Binary | Intracephalic hemorrhage (ICD I61) |
| | subsequent_myocardial_infarction | Binary | Subsequent myocardial infarction (ICD I22) |
| | acute_pericarditis | Binary | Acute pericarditis (ICD I30) |
| | post_procedural_disorders_of_the_circulatory_system_not_elsewhere_classified | Binary | Post procedural disorders of the circulatory system not elsewhere classified (ICD I97) |
| | other_acute_ischemic_heart_diseases | Binary | Other acute ischemic heart diseases (ICD I24) |
| | hypertensive_cardiorenal_disease | Binary | Hypertensive cardiorenal disease (ICD I13) |
| | subarachnoid_hemorrhage | Binary | Subarachnoid hemorrhage (ICD I60) |
| | pulmonary_valve_disorders | Binary | Pulmonary valve disorders (ICD I37) |
| | other_non_traumatic_intracranial_hemorrhages | Binary | Other non-traumatic intracranial hemorrhages (ICD I62) |
| | acute_and_subacute_endocarditis | Binary | Acute and subacute endocarditis (ICD I33) |
| | certain_complications_present_after_acute_myocardial_infarction | Binary | Certain complications present after acute myocardial infarction (ICD I23) |
| | portal_vein_thrombosis | Binary | Portal vein thrombosis (ICD I81) |
| | rheumatic_fever_without_mention_of_cardiac_complication | Binary | Rheumatic fever without mention of cardiac complication (ICD I00) |
| | nonrheumatic_disorders_of_the_tricuspid_valve | Binary | Nonrheumatic disorders of the tricuspid valve (ICD I36) |
| | disorders_of_lipoprotein_metabolism_and_other_lipidemias | Binary | Disorders of lipoprotein metabolism and other lipidemias (ICD E78) |
| | obesity | Binary | Obesity (ICD E66) |
| | diabetes_mellitus_unspecified | Binary | Unspecified diabetes mellitus (ICD E14) |
| | other_hypothyroidisms | Binary | Other hypothyroidisms (ICD E03) |
| | type_1_diabetes_mellitus | Binary | Type 1 diabetes mellitus (ICD E10) |
| | other_disorders_of_carbohydrate_metabolism | Binary | Other disorders of carbohydrate metabolism (ICD E74) |
| | other_metabolic_disorders | Binary | Other metabolic disorders (ICD E88) |
| | purine_and_pyrimidine_metabolism_disorders | Binary | Purine and pyrimidine metabolism disorders (ICD E79) |
| | hypoparathyroidism | Binary | Hypoparathyroidism (ICD E20) |

| | | | |
|---|---|---|---|
| | post_procedural_endocrine_and_metabolic_disorders_not_elsewhere_classified | Binary | Post procedural endocrine and metabolic disorders not elsewhere classified (ICD E89) |
| | hyperfunction_of_the_pituitary_gland | Binary | Hyperfunction of the pituitary gland (ICD E22) |
| | other_disorders_of_the_adrenal_gland | Binary | Other disorders of the adrenal gland (ICD E27) |
| | other_non_toxic_goiters | Binary | Other non-toxic goiters (ICD E04) |
| | hypofunction_and_other_disorders_of_the_pituitary_gland | Binary | Hypofunction and other disorders of the pituitary gland (ICD E23) |
| | thyrotoxicosis_hyperthyroidism | Binary | Hyperthyroidism thyrotoxicosis (ICD E05) |
| | other_specified_diabetes_mellitus | Binary | Other specified diabetes mellitus (ICD E13) |
| | ovarian_dysfunction | Binary | Ovarian dysfunction (ICD E28) |
| | deficiencies_of_other_nutritional_elements | Binary | Deficiencies of other nutritional elements (ICD E61) |
| | other_endocrine_disorders | Binary | Other endocrine disorders (ICD E34) |
| | other_thyroid_disorders | Binary | Other thyroid disorders (ICD E07) |
| | lactose_intolerance | Binary | Lactose intolerance (ICD E73) |
| | moderate_and_mild_protein_calorie_malnutrition | Binary | Moderate and mild protein-calorie malnutrition (ICD E44) |
| | protein_calorie_malnutrition_unspecified | Binary | Unspecified protein-calorie malnutrition (ICD E46) |
| | severe_protein_calorie_malnutrition_unspecified | Binary | Unspecified severe protein-calorie malnutrition (ICD E43) |
| | other_fluid_electrolyte_and_acid_base_disorders | Binary | Other disorders of fluid electrolytes and acid-base balance (ICD E87) |
| | diabetes_mellitus_associated_with_malnutrition | Binary | Diabetes mellitus associated with malnutrition (ICD E12) |
| | vitamin_a_deficiency | Binary | Vitamin A deficiency (ICD E50) |
| | hyperparathyroidism_and_other_disorders_of_the_parathyroid_gland | Binary | Hyperparathyroidism and other disorders of the parathyroid gland (ICD E21) |
| | mineral_metabolism_disorders | Binary | Mineral metabolism disorders (ICD E83) |
| | other_vitamin_deficiencies | Binary | Other vitamin deficiencies (ICD E56) |
| | nutritional_marasmus | Binary | Nutritional marasmus (ICD E41) |
| | amyloidosis | Binary | Amyloidosis (ICD E85) |
| | thyroiditis | Binary | Thyroiditis (ICD E06) |
| | volume_depletion | Binary | Volume depletion (ICD E86) |

| | | | |
|---|---|---|---|
| | thyroid_disorders_linked_to_iodine_deficiency_and_related_conditions | Binary | Thyroid disorders linked to iodine deficiency and related conditions (ICD E01) |
| | subclinical_hypothyroidism_due_to_iodine_deficiency | Binary | Subclinical hypothyroidism due to iodine deficiency (ICD E02) |
| | cystic_fibrosis | Binary | Cystic fibrosis (ICD E84) |
| | other_disorders_of_the_internal_secretion_of_the_pancreas | Binary | Other disorders of the internal secretion of the pancreas (ICD E16) |
| | cushing's_syndrome | Binary | Cushing's syndrome (ICD E24) |
| | testicular_dysfunction | Binary | Testicular dysfunction (ICD E29) |
| | hyperaldosteronism | Binary | Hyperaldosteronism (ICD E26) |
| | deficiencies_of_other_b_vitamins | Binary | Deficiencies of other B vitamins (ICD E53) |
| | dietary_calcium_deficiency | Binary | Dietary calcium deficiency (ICD E58) |
| | disorders_of_sphingolipid_metabolism_and_other_lipid_storage_disorders | Binary | Disorders of sphingolipid metabolism and other lipid storage disorders (ICD E75) |
| | developmental_delay_due_to_protein_calorie_malnutrition | Binary | Developmental delay due to protein-calorie malnutrition (ICD E45) |
| | aromatic_amino_acid_metabolism_disorders | Binary | Aromatic amino acid metabolism disorders (ICD E70) |
| | disorders_of_porphyrin_and_bilirubin_metabolism | Binary | Disorders of porphyrin and bilirubin metabolism (ICD E80) |
| | localized_adiposity | Binary | Localized adiposity (ICD E65) |
| | sequelae_of_malnutrition_and_other_nutritional_deficiencies | Binary | Sequelae of malnutrition and other nutritional deficiencies (ICD E64) |
| | vitamin_d_deficiency | Binary | Vitamin D deficiency (ICD E55) |
| | non_diabetic_hypoglycemic_coma | Binary | Non-diabetic hypoglycemic coma (ICD E15) |
| | other_nutritional_deficiencies | Binary | Other types of hyperalimentation (ICD E67) |
| | diseases_of_the_thymus | Binary | Other nutritional deficiencies (ICD E63) |
| | disorders_of_the_metabolism_of_branched_chain_amino_acids_and_fatty_acids | Binary | Disorders of metabolism of branched-chain amino acids and fatty acids (ICD E71) |
| | other_disorders_of_amino_acid_metabolism | Binary | Other disorders of amino acid metabolism (ICD E72) |
| | marasmatic_kwashiorkor | Binary | Marasmic kwashiorkor (ICD E42) |
| | niacin_deficiency_pellagra | Binary | Pellagra niacin deficiency (ICD E52) |
| | polyglandular_dysfunction | Binary | Polyglandular dysfunction (ICD E31) |
| | ascorbic_acid_deficiency | Binary | Ascorbic acid deficiency (ICD E54) |
| | adrenogenital_disorders | Binary | Adrenogenital disorders (ICD E25) |

| | | | |
|---|---|---|---|
| | glycosaminoglycan_metabolism_disorders | Binary | Glycosaminoglycan metabolism disorders (ICD E76) |
| | congenital_iodine_deficiency_syndrome | Binary | Congenital iodine deficiency syndrome (ICD E00) |
| | type_2_diabetes_mellitus_without_mention_of_complication | Binary | Type 2 diabetes mellitus without mention of complication (ICD E11.9) |
| | type_2_diabetes_mellitus_with_other_specified_complications | Binary | Type 2 diabetes mellitus with other specified complications (ICD E11.6) |
| | type_2_diabetes_mellitus_with_multiple_complications | Binary | Type 2 diabetes mellitus with multiple complications (ICD E11.7) |
| | type_2_diabetes_mellitus_with_ophthalmic_complications | Binary | Type 2 diabetes mellitus with ophthalmic complications (ICD E11.3) |
| | type_2_diabetes_mellitus_with_peripheral_circulatory_complications | Binary | Type 2 diabetes mellitus with peripheral circulatory complications (ICD E11.5) |
| | type_2_diabetes_mellitus_with_unspecified_complications | Binary | Type 2 diabetes mellitus with unspecified complications (ICD E11.8) |
| | type_2_diabetes_mellitus_with_renal_complications | Binary | Type 2 diabetes mellitus with renal complications (ICD E11.2) |
| | type_2_diabetes_mellitus_with_neurological_complications | Binary | Type 2 diabetes mellitus with neurological complications (ICD E11.4) |
| | diabetes_mellitus_type_2_with_coma | Binary | Type 2 diabetes mellitus with coma (ICD E11.0) |
| | type_2_diabetes_mellitus_with_ketoacidosis | Binary | Type 2 diabetes mellitus with ketoacidosis (ICD E11.1) |
| Body measurements (45) | fn_weight_mean | Float | Mean Weight (kg) value |
| | bmi_value | Float | Value of BMI (kg/m2) value |
| | bmi_label | String | Group of BMI (kg/m2) value |
| | bmi_ordinal | Integer | Ordinal group of BMI (kg/m2) value |
| | fn_weight_median | Float | Median Weight (kg) value |
| | fn_weight_max | Float | Max Weight (kg) value |
| | fn_weight_min | Float | Min Weight (kg) value |
| | fn_weight_std | Float | Standard deviation Weight (kg) value |
| | fn_weight_slope | Float | Slope Weight (kg) value |
| | fn_height_mean | Float | Mean Height (m) value |
| | fn_height_median | Float | Median Height (m) value |
| | fn_height_max | Float | Max Height (m) value |
| | fn_height_min | Float | Min Height (m) value |
| | fn_height_std | Float | Standard deviation Height (m) value |
| | fn_height_slope | Float | Slope Height (m) value |

| | | | |
|---|---|---|---|
| | fn_ta_systolic_mean | Float | Mean Systolic blood pressure (mmHg) value |
| | fn_ta_systolic_median | Float | Median Systolic blood pressure (mmHg) value |
| | fn_ta_systolic_max | Float | Max Systolic blood pressure (mmHg) value |
| | fn_ta_systolic_min | Float | Min Systolic blood pressure (mmHg) value |
| | fn_ta_systolic_std | Float | Standard deviation Systolic blood pressure (mmHg) value |
| | fn_ta_systolic_slope | Float | Slope Systolic blood pressure (mmHg) value |
| | fn_ta_diastolic_mean | Float | Mean Diastolic blood pressure (mmHg) value |
| | fn_ta_diastolic_median | Float | Median Diastolic blood pressure (mmHg) value |
| | fn_ta_diastolic_max | Float | Max Diastolic blood pressure (mmHg) value |
| | fn_ta_diastolic_min | Float | Min Diastolic blood pressure (mmHg) value |
| | fn_ta_diastolic_std | Float | Standard deviation Diastolic blood pressure (mmHg) value |
| | fn_ta_diastolic_slope | Float | Slope Diastolic blood pressure (mmHg) value |
| | in_heart_rate_mean | Float | Mean Heart rate ([U]/min) value |
| | in_heart_rate_median | Float | Median Heart rate ([U]/min) value |
| | in_heart_rate_max | Float | Max Heart rate ([U]/min) value |
| | in_heart_rate_min | Float | Min Heart rate ([U]/min) value |
| | in_heart_rate_std | Float | Standard deviation Heart rate ([U]/min) value |
| | in_heart_rate_slope | Float | Slope Heart rate ([U]/min) value |
| | in_respiratory_frequency_mean | Float | Mean Breath rate ([U]/min) value |
| | in_respiratory_frequency_median | Float | Median Breath rate ([U]/min) value |
| | in_respiratory_frequency_max | Float | Max Breath rate ([U]/min) value |
| | in_respiratory_frequency_min | Float | Min Breath rate ([U]/min) value |
| | in_respiratory_frequency_std | Float | Standard deviation Breath rate ([U]/min) value |
| | in_respiratory_frequency_slope | Float | Slope Breath rate ([U]/min) value |
| | fn_temperature_mean | Float | Mean Temperature (ºC) value |
| | fn_temperature_median | Float | Median Temperature (ºC) value |
| | fn_temperature_max | Float | Max Temperature (ºC) value |
| | fn_temperature_min | Float | Min Temperature (ºC) value |
| | fn_temperature_std | Float | Standard deviation Temperature (ºC) value |
| | fn_temperature_slope | Float | Slope Temperature (ºC) value |
| Laboratory results (129) | in_glucose_mean | Float | Mean Glucose (mg/dL) value |
| | glucose_value | Float | Value of Glucose (mg/dL) value |

| | | | |
|---|---|---|---|
| | glucose_label | String | Group of Glucose (mg/dL) value |
| | glucose_ordinal | Integer | Ordinal group of Glucose (mg/dL) value |
| | in_glucose_median | Float | Median Glucose (mg/dL) value |
| | in_glucose_max | Float | Max Glucose (mg/dL) value |
| | in_glucose_min | Float | Min Glucose (mg/dL) value |
| | in_glucose_std | Float | Standard deviation Glucose (mg/dL) value |
| | in_glucose_slope | Float | Slope Glucose (mg/dL) value |
| | fn_fasting_glucose_mean | Float | Mean Fasting glucose (mg/dL) value |
| | fn_fasting_glucose_median | Float | Median Fasting glucose (mg/dL) value |
| | fn_fasting_glucose_max | Float | Max Fasting glucose (mg/dL) value |
| | fn_fasting_glucose_min | Float | Min Fasting glucose (mg/dL) value |
| | fn_fasting_glucose_std | Float | Standard deviation Fasting glucose (mg/dL) value |
| | fn_fasting_glucose_slope | Float | Slope Fasting glucose (mg/dL) value |
| | fn_capillary_glucose_mean | Float | Mean Capillary glucose (mg/dL) value |
| | fn_capillary_glucose_median | Float | Median Capillary glucose (mg/dL) value |
| | fn_capillary_glucose_max | Float | Max Capillary glucose (mg/dL) value |
| | fn_capillary_glucose_min | Float | Min Capillary glucose (mg/dL) value |
| | fn_capillary_glucose_std | Float | Standard deviation Capillary glucose (mg/dL) value |
| | fn_capillary_glucose_slope | Float | Slope Capillary glucose (mg/dL) value |
| | fn_hemoglobin_mean | Float | Mean HbA1c (%) value |
| | hemoglobin_value | Float | Value of HbA1c (%) value |
| | hemoglobin_label | String | Group of HbA1c (%) value |
| | hemoglobin_ordinal | Integer | Ordinal group of HbA1c (%) value |
| | fn_hemoglobin_median | Float | Median HbA1c (%) value |
| | fn_hemoglobin_max | Float | Max HbA1c (%) value |
| | fn_hemoglobin_min | Float | Min HbA1c (%) value |
| | fn_hemoglobin_std | Float | Standard deviation HbA1c (%) value |
| | fn_hemoglobin_slope | Float | Slope HbA1c (%) value |
| | fn_triglycerides_mean | Float | Mean Triglycerides (mg/dL) value |
| | triglycerides_value | Float | Value of Triglycerides (mg/dL) value |
| | triglycerides_label | String | Group of Triglycerides (mg/dL) value |
| | triglycerides_ordinal | Integer | Ordinal group of Triglycerides (mg/dL) value |
| | fn_triglycerides_median | Float | Median Triglycerides (mg/dL) value |
| | fn_triglycerides_max | Float | Max Triglycerides (mg/dL) value |
| | fn_triglycerides_min | Float | Min Triglycerides (mg/dL) value |
| | fn_triglycerides_std | Float | Standard deviation Triglycerides (mg/dL) value |

| | | | |
|---|---|---|---|
| | fn_triglycerides_slope | Float | Slope Triglycerides (mg/dL) value |
| | fn_right_foot_mean | Float | Mean Diabetic right foot value |
| | fn_right_foot_median | Float | Median Diabetic right foot value |
| | fn_right_foot_max | Float | Max Diabetic right foot value |
| | fn_right_foot_min | Float | Min Diabetic right foot value |
| | fn_right_foot_std | Float | Standard deviation Diabetic right foot value |
| | fn_right_foot_slope | Float | Slope Diabetic right foot value |
| | fn_left_foot_mean | Float | Mean Diabetic left foot value |
| | diabetic_right_foot_ordinal | Integer | Ordinal Diabetic right foot value |
| | diabetic_left_foot_ordinal | Integer | Ordinal Diabetic left foot value |
| | fn_left_foot_median | Float | Median Diabetic left foot value |
| | fn_left_foot_max | Float | Max Diabetic left foot value |
| | fn_left_foot_min | Float | Min Diabetic left foot value |
| | fn_left_foot_std | Float | Standard deviation Diabetic left foot value |
| | fn_left_foot_slope | Float | Slope Diabetic left foot value |
| | fn_ph_mean | Float | Mean Hydrogen potential in urine value |
| | fn_ph_median | Float | Median Hydrogen potential in urine value |
| | fn_ph_max | Float | Max Hydrogen potential in urine value |
| | fn_ph_min | Float | Min Hydrogen potential in urine value |
| | fn_ph_std | Float | Standard deviation Hydrogen potential in urine value |
| | fn_ph_slope | Float | Slope Hydrogen potential in urine value |
| | fn_density_mean | Float | Mean Urine density (g/mL) value |
| | fn_density_median | Float | Median Urine density (g/mL) value |
| | fn_density_max | Float | Max Urine density (g/mL) value |
| | fn_density_min | Float | Min Urine density (g/mL) value |
| | fn_density_std | Float | Standard deviation Urine density (g/mL) value |
| | fn_density_slope | Float | Slope Urine density (g/mL) value |
| | fn_ego_mean | Float | Mean General urine test value |
| | fn_ego_median | Float | Median General urine test value |
| | fn_ego_max | Float | Max General urine test value |
| | fn_ego_min | Float | Min General urine test value |
| | fn_ego_std | Float | Standard deviation General urine test value |
| | fn_ego_slope | Float | Slope General urine test value |
| | fn_urea_mean | Float | Mean Urea in urine (mg/dL) value |
| | fn_urea_median | Float | Median Urea in urine (mg/dL) value |

| | | | |
|---|---|---|---|
| | fn_urea_max | Float | Max Urea in urine (mg/dL) value |
| | fn_urea_min | Float | Min Urea in urine (mg/dL) value |
| | fn_urea_std | Float | Standard deviation Urea in urine (mg/dL) value |
| | fn_urea_slope | Float | Slope Urea in urine (mg/dL) value |
| | fn_glycemia_mean | Float | Mean Blood glucose (mg/dL) value |
| | fn_glycemia_median | Float | Median Blood glucose (mg/dL) value |
| | fn_glycemia_max | Float | Max Blood glucose (mg/dL) value |
| | fn_glycemia_min | Float | Min Blood glucose (mg/dL) value |
| | fn_glycemia_std | Float | Standard deviation Blood glucose (mg/dL) value |
| | fn_glycemia_slope | Float | Slope Blood glucose (mg/dL) value |
| | fn_aurico_mean | Float | Mean Uric acid in urine (mg/dL) value |
| | fn_aurico_median | Float | Median Uric acid in urine (mg/dL) value |
| | fn_aurico_max | Float | Max Uric acid in urine (mg/dL) value |
| | fn_aurico_min | Float | Min Uric acid in urine (mg/dL) value |
| | fn_aurico_std | Float | Standard deviation Uric acid in urine (mg/dL) value |
| | fn_aurico_slope | Float | Slope Uric acid in urine (mg/dL) value |
| | fn_creatinine_mean | Float | Mean Creatinine (mg/dL) value |
| | gfr_value | Float | Value of Glomerular filtration rate (mg/dL) value |
| | gfr_label | String | Group of Glomerular filtration rate (mg/dL) value |
| | gfr_ordinal | Integer | Ordinal group of Glomerular filtration rate (mg/dL) value |
| | creatinine_value | Float | Value of Creatinine (mg/dL) value |
| | creatinine_label | String | Group of Creatinine (mg/dL) value |
| | creatinine_ordinal | Integer | Ordinal group of Creatinine (mg/dL) value |
| | fn_creatinine_median | Float | Median Creatinine (mg/dL) value |
| | fn_creatinine_max | Float | Max Creatinine (mg/dL) value |
| | fn_creatinine_min | Float | Min Creatinine (mg/dL) value |
| | fn_creatinine_std | Float | Standard deviation Creatinine (mg/dL) value |
| | fn_creatinine_slope | Float | Slope Creatinine (mg/dL) value |
| | fn_ego_density_mean | Float | Mean General urine test density (g/mL) value |
| | fn_ego_density_median | Float | Median General urine test density (g/mL) value |
| | fn_ego_density_max | Float | Max General urine test density (g/mL) value |
| | fn_ego_density_min | Float | Min General urine test density (g/mL) value |
| | fn_ego_density_std | Float | Standard deviation General urine test density (g/mL) value |
| | fn_ego_density_slope | Float | Slope General urine test density (g/mL) value |

| | | | |
|---|---|---|---|
| | fn_urine_culture_mean | Float | Mean Urine culture value |
| | fn_urine_culture_median | Float | Median Urine culture value |
| | fn_urine_culture_max | Float | Max Urine culture value |
| | fn_urine_culture_min | Float | Min Urine culture value |
| | fn_cholesterol_mean | Float | Mean Cholesterol (mg/dL) value |
| | cholesterol_value | Float | Value of Cholesterol (mg/dL) value |
| | cholesterol_label | String | Group of Cholesterol (mg/dL) value |
| | cholesterol_ordinal | Integer | Ordinal group of Cholesterol (mg/dL) value |
| | fn_cholesterol_median | Float | Median Cholesterol (mg/dL) value |
| | fn_cholesterol_max | Float | Max Cholesterol (mg/dL) value |
| | fn_cholesterol_min | Float | Min Cholesterol (mg/dL) value |
| | fn_cholesterol_std | Float | Standard deviation Cholesterol (mg/dL) value |
| | fn_cholesterol_slope | Float | Slope Cholesterol (mg/dL) value |
| | albumin | Binary | Albumin presence in urine |
| | bacteria | Binary | Bacteria presence in urine |
| | acetone_bilirubin | Binary | Acetone bilirubin presence in urine |
| | cylinders | Binary | Cylinders presence in urine |
| | erythrocytes | Binary | Erythrocytes presence in urine |
| | glucose | Binary | Glucose presence in urine |
| | hemoglobin | Binary | Hemoglobin presence in urine |
| | leukocytes | Binary | Leukocytes presence in urine |
| | others | Binary | Others presences in urine |
| Drugs (110) | antivertigious_sum | Float | Sum of prescribed Antivertiginous drug dose |
| | antivertigious_mean | Float | Mean of prescribed Antivertiginous drug dose |
| | antivertigious_slope | Float | Slope of prescribed Antivertiginous drug dose |
| | antihypertensives_sum | Float | Sum of prescribed Antihypertensives drug dose |
| | antihypertensives_mean | Float | Mean of prescribed Antihypertensives drug dose |
| | antihypertensives_slope | Float | Slope of prescribed Antihypertensives drug dose |
| | antiuricosuricos_sum | Float | Sum of prescribed Anti Uricosurics drug dose |
| | antiuricosuricos_mean | Float | Mean of prescribed Anti Uricosurics drug dose |
| | antiuricosuricos_slope | Float | Slope of prescribed Anti Uricosurics drug dose |
| | bronchodilators_and_expectorants_sum | Float | Sum of prescribed Bronchodilators and expectorants drug dose |
| | bronchodilators_and_expectorants_mean | Float | Mean of prescribed Bronchodilators and expectorants drug dose |
| | bronchodilators_and_expectorants_slope | Float | Slope of prescribed Bronchodilators and expectorants drug dose |

| | | | |
|---|---|---|---|
| | antiulcerous_and_protectors_of_the_gastric_mucosa_sum | Float | Sum of prescribed Antiulceratives and protectors of the gastric mucosa drug dose |
| | antiulcerous_and_protectors_of_the_gastric_mucosa_mean | Float | Mean of prescribed Antiulceratives and protectors of the gastric mucosa drug dose |
| | antiulcerous_and_protectors_of_the_gastric_mucosa_slope | Float | Slope of prescribed Antiulceratives and protectors of the gastric mucosa drug dose |
| | ocular_drugs_sum | Float | Sum of prescribed Ocular pharmaceuticals drug dose |
| | ocular_drugs_mean | Float | Mean of prescribed Ocular pharmaceuticals drug dose |
| | ocular_drugs_slope | Float | Slope of prescribed Ocular pharmaceuticals drug dose |
| | antidiabetics_sum | Float | Sum of prescribed Antidiabetic drug dose |
| | antidiabetics_mean | Float | Mean of prescribed Antidiabetic drug dose |
| | antidiabetics_slope | Float | Slope of prescribed Antidiabetic drug dose |
| | antidepressants_sum | Float | Sum of prescribed Antidepressants drug dose |
| | antidepressants_mean | Float | Mean of prescribed Antidepressants drug dose |
| | antidepressants_slope | Float | Slope of prescribed Antidepressants drug dose |
| | anti_inflammatory_non_steroids_sum | Float | Sum of prescribed Non-steroidal anti-inflammatory drugs drug dose |
| | anti_inflammatory_non_steroids_mean | Float | Mean of prescribed Non-steroidal anti-inflammatory drugs drug dose |
| | anti_inflammatory_non_steroid_slope | Float | Slope of prescribed Non-steroidal anti-inflammatory drugs drug dose |
| | antibiotics_sum | Float | Sum of prescribed Antibiotics drug dose |
| | antibiotics_mean | Float | Mean of prescribed Antibiotics drug dose |
| | antibiotics_slope | Float | Slope of prescribed Antibiotics drug dose |
| | hormonal_sum | Float | Sum of prescribed Hormonal drug dose |
| | hormonal_mean | Float | Mean of prescribed Hormonal drug dose |
| | hormonal_slope | Float | Slope of prescribed Hormonal drug dose |
| | vitamins_sum | Float | Sum of prescribed Vitamins drug dose |
| | vitamins_mean | Float | Mean of prescribed Vitamins drug dose |
| | vitamins_slope | Float | Slope of prescribed Vitamins drug dose |
| | antitussives_sum | Float | Sum of prescribed Antitussives drug dose |
| | antitussives_mean | Float | Mean of prescribed Antitussives drug dose |
| | antitussives_slope | Float | Slope of prescribed Antitussives drug dose |
| | anti_allergic_sum | Float | Sum of prescribed Antiallergic drug dose |
| | anti_allergic_mean | Float | Mean of prescribed Antiallergic drug dose |
| | anti_allergic_slope | Float | Slope of prescribed Antiallergic drug dose |

| | | | |
|---|---|---|---|
| | antiplatelets_sum | Float | Sum of prescribed Antiplatelet agents drug dose |
| | antiplatelets_mean | Float | Mean of prescribed Antiplatelet agents drug dose |
| | antiplatelets_slope | Float | Slope of prescribed Antiplatelet agents drug dose |
| | oral_electrolytes_sum | Float | Sum of prescribed Oral electrolytes drug dose |
| | oral_electrolytes_mean | Float | Mean of prescribed Oral electrolytes drug dose |
| | oral_electrolytes_slope | Float | Slope of prescribed Oral electrolytes drug dose |
| | antineuritics_sum | Float | Sum of prescribed Antineuritics drug dose |
| | antineuritics_mean | Float | Mean of prescribed Antineuritics drug dose |
| | antineuritics_slope | Float | Slope of prescribed Antineuritics drug dose |
| | methylxanthines_sum | Float | Sum of prescribed Methylxanthines drug dose |
| | methylxanthines_mean | Float | Mean of prescribed Methylxanthines drug dose |
| | methylxanthines_slope | Float | Slope of prescribed Methylxanthines drug dose |
| | disease_modifiing_drugs_sum | Float | Sum of prescribed Disease modifying drugs drug dose |
| | disease_modifiing_drugs_mean | Float | Mean of prescribed Disease modifying drugs drug dose |
| | disease_modifiing_drugs_slope | Float | Slope of prescribed Disease modifying drugs drug dose |
| | drugs_used_in_nephrology_sum | Float | Sum of prescribed Drugs used in nephrology drug dose |
| | drugs_used_in_nephrology_mean | Float | Mean of prescribed Drugs used in nephrology drug dose |
| | drugs_used_in_nephrology_slope | Float | Slope of prescribed Drugs used in nephrology drug dose |
| | analgesics_urinary_sum | Float | Sum of prescribed Urinary analgesics drug dose |
| | analgesics_urinary_mean | Float | Mean of prescribed Urinary analgesics drug dose |
| | analgesics_urinary_slope | Float | Slope of prescribed Urinary analgesics drug dose |
| | benzodiazepines_sum | Float | Sum of prescribed Benzodiazepines drug dose |
| | benzodiazepines_mean | Float | Mean of prescribed Benzodiazepines drug dose |
| | benzodiazepines_slope | Float | Slope of prescribed Benzodiazepines drug dose |
| | systemic_and_topical_antifungals_sum | Float | Sum of prescribed Systemic and topical Antifungals drug dose |
| | systemic_and_topical_antifungals_mean | Float | Mean of prescribed Systemic and topical Antifungals drug dose |
| | systemic_and_topical_antifungals_slope | Float | Slope of prescribed Systemic and topical Antifungals drug dose |
| | lipid_lowering_sum | Float | Sum of prescribed Lipid-lowering drug dose |
| | lipid_lowering_mean | Float | Mean of prescribed Lipid-lowering drug dose |
| | lipid_lower_slope | Float | Slope of prescribed Lipid-lowering drug dose |

| | | | |
|---|---|---|---|
| | drugs_used_in_neurology_sum | Float | Sum of prescribed Drugs used in neurology drug dose |
| | drugs_used_in_neurology_mean | Float | Mean of prescribed Drugs used in neurology drug dose |
| | drugs_used_in_neurology_slope | Float | Slope of prescribed Drugs used in neurology drug dose |
| | antiflamatory_steroids_sum | Float | Sum of prescribed Steroidal anti-inflammatories drug dose |
| | antiflamatory_steroids_mean | Float | Mean of prescribed Steroidal anti-inflammatories drug dose |
| | antiflamatory_steroids_slope | Float | Slope of prescribed Steroidal anti-inflammatories drug dose |
| | alpha_blockers_sum | Float | Sum of prescribed Alpha blockers drug dose |
| | alpha_blockers_mean | Float | Mean of prescribed Alpha blockers drug dose |
| | alpha_blockers_slope | Float | Slope of prescribed Alpha blockers drug dose |
| | drugs_used_in_psychiatry_sum | Float | Sum of prescribed Drugs used in psychiatry drug dose |
| | drugs_used_in_psychiatry_mean | Float | Mean of prescribed Drugs used in psychiatry drug dose |
| | drugs_used_in_psychiatry_slope | Float | Slope of prescribed Drugs used in psychiatry drug dose |
| | erectile_dysfunction_sum | Float | Sum of prescribed Erectile dysfunction drug dose |
| | erectile_dysfunction_mean | Float | Mean of prescribed Erectile dysfunction drug dose |
| | erectile_dysfunction_slope | Float | Slope of prescribed Erectile dysfunction drug dose |
| | drugs_used_in_hypothyroidism_sum | Float | Sum of prescribed Drugs used in hypothyroidism drug dose |
| | drugs_used_in_hypothyroidism_mean | Float | Mean of prescribed Drugs used in hypothyroidism drug dose |
| | drugs_used_in_hypothyroidism_slope | Float | Slope of prescribed Drugs used in hypothyroidism drug dose |
| | antimuscarinics_sum | Float | Sum of prescribed Antimuscarinics drug dose |
| | antimuscarinics_mean | Float | Mean of prescribed Antimuscarinics drug dose |
| | antimuscarinics_slope | Float | Slope of prescribed Antimuscarinics drug dose |
| | antiandrogenic_sum | Float | Sum of prescribed Antiandrogenic drug dose |
| | antiandrogenic_mean | Float | Mean of prescribed Antiandrogenic drug dose |
| | antiandrogenic_slope | Float | Slope of prescribed Antiandrogenic drug dose |
| | drugs_used_in_hyperthyroidism_sum | Float | Sum of prescribed Drugs used in hyperthyroidism drug dose |
| | drugs_used_in_hyperthyroidism_mean | Float | Mean of prescribed Drugs used in hyperthyroidism drug dose |
| | drugs_used_in_hyperthyroidism_slope | Float | Slope of prescribed Drugs used in hyperthyroidism drug dose |

| | | | |
|---|---|---|---|
| | complete_nutritional_formulas_sum | Float | Sum of prescribed Complete nutritional formulas drug dose |
| | complete_nutritional_formulas_mean | Float | Mean of prescribed Complete nutritional formulas drug dose |
| | complete_nutritional_formulas_slope | Float | Slope of prescribed Complete nutritional formulas drug dose |
| | drugs_used_in_breast_cancer_sum | Float | Sum of prescribed Drugs used in breast cancer drug dose |
| | drugs_used_in_breast_cancer_mean | Float | Mean of prescribed Drugs used in breast cancer drug dose |
| | drugs_used_in_breast_cancer_slope | Float | Slope of prescribed Drugs used in breast cancer drug dose |
| | drugs_used_in_osteoporosis_sum | Float | Sum of prescribed Drugs used in osteoporosis drug dose |
| | drugs_used_in_osteoporosis_mean | Float | Mean of prescribed Drugs used in osteoporosis drug dose |
| | drugs_used_in_osteoporosis_slope | Float | Slope of prescribed Drugs used in osteoporosis drug dose |
| | antiarrhythmics_sum | Float | Sum of prescribed Antiarrhythmic drug dose |
| | antiarrhythmics_mean | Float | Mean of prescribed Antiarrhythmic drug dose |
| Predictions (5) | e11 | Integer | Type 2 diabetes mellitus (ICD E11) diagnosis in next year |
| | e112 | Integer | Type 2 diabetes mellitus with kidney complications (ICD E11.2) diagnosis in next year |
| | e113 | Integer | Type 2 diabetes mellitus with ophthalmic complications (ICD E11.3) diagnosis in next year |
| | e114 | Integer | Type 2 diabetes mellitus with neurological complications (ICD E11.4) diagnosis in next year |
| | e115 | Integer | Type 2 diabetes mellitus with peripheral circulatory complications (ICD E11.5) diagnosis in next year |

#### F. Categorical Variables

To determine categorical variables, previously established data were taken into consideration for variables such as Body Mass Index, triglycerides and cholesterol classification, as well as the classification of renal insufficiency (Creatinine classification), which are previously described and accepted by international consensus.

The following categories were established for each variable,

For Body Mass Index, the following was considered: BMI classifications are based on cardiovascular disease risk. These BMI classifications have been adopted by NIH and WHO for white, Hispanic, and black individuals. Two categories are defined for BMI: To determine the ranges of body mass index, all those subjects who met a body mass index <24.9 were taken into account to grant the normal category and the remaining subjects were categorized as Overweight & Obesity, thus respecting a binarization of variables for effective interpretation.

Body mass index (BMI) classification:

<table>
<tr><th>Range</th><th>Classification</th><th>Binary Categories</th></tr>
<tr><td>< 18.5 kg/m2</td><td>Underweight</td><td rowspan="2">Normal</td></tr>
<tr><td>18.5 kg/m2 - 24.9 kg/m2</td><td>Normal</td></tr>
<tr><td>25 kg/m2 - 29.9 kg/m2</td><td>Overweight</td><td rowspan="4">Overweight & Obesity</td></tr>
<tr><td>30 kg/m2 - 34.9 kg/m2</td><td>Obesity (class 1)</td></tr>
<tr><td>35 kg/m2 - 39.9 kg/m2</td><td>Obesity (class 2)</td></tr>
<tr><td>40.0 kg/m2 ></td><td>Obesity (class 3)</td></tr>
</table>

To determine the ranges of glucose and glycated hemoglobin, the distribution of the patient population in the source database was taken into consideration in order to standardize the measurements and harmonize the selected quintiles. The proportions of the distributions are shown in the graph below, and the cutoff values were assigned according to their distribution and do not follow a previously selected standard. Finally, a binarization of the variable was assigned to establish an adequate interpretation by establishing a normal value <125 mg/dl and a high value starting from 126 mg/dl and up to a value > 450 mg/dl.

Glucose classification:

To determine the ranges of glucose and glycated hemoglobin, we use the standards of care for diabetes as outlined in the Diabetes Classification and Diagnosis Guideline: 2023[1] were taken into consideration. The distribution of the patient population in the source database was taken into consideration in order to standardize the measurements and harmonize the selected quintiles. The proportions of the distributions are shown in the graph below, and the cutoff values were assigned according to their distribution and do not follow a previously selected standard. Finally, a binarization of the variable was assigned to establish an adequate interpretation by establishing a normal value <125 mg/dl and a high value starting from 126 mg/dl and up to a value > 450 mg/dl.

| Range | Category | Binary |
|---|---|---|
| 25 mg/dL - 59 mg/dL | A | Normal |
| 60 mg/dL - 125 mg/dL | B | |
| 126 mg/dL - 249 mg/dL | C | High |
| 250 mg/dL - 449 mg/dL | D | |
| > 450 mg/dL | E | |

Normal HbA1c values: For people without diabetes, the normal range for HbA1c is usually below 5.7%. Abnormal HbA1c values:In people with diabetes, HbA1c control goals may vary based on medical guidelines, but in general, the following is considered: HbA1c less than 6.5%: For this reason, it was adjusted that the population with a glycosylated hemoglobin greater than 6 and up to a value greater than 17.3 would be categorized into a binary variable as a high value of glycosylated hemoglobin.

Hemoglobin HbA1c classification[2]:

| Range | Category | Binary |
|---|---|---|
| 2.5 % - 3.69 % | A | Normal |
| 3.7 % - 5.99 % | B | |

[1] https://diabetesjournals.org/care/article/46/Supplement_1/S19/148056/2-Classification-and-Diagnosis-of-Diabetes
[2] https://diabetesjournals.org/care/article/46/Supplement_1/S19/148056/2-Classification-and-Diagnosis-of-Diabetes

| | | |
|---|---|---|
| 6 % - 10.29 % | C | High |
| 10.3 % - 17.29 % | D | |
| > 17.3 % | E | |

For triglycerides, values were classified based on whether patients had normal triglyceridemia or moderate, moderate to severe, or severe hypertriglyceridemia. Normal triglyceridemia was defined as triglyceride levels < 150 mg/dL (< 1.7 mmol/L),Triglycerides classification[3]: For this reason, it was adjusted that the population with triglyceride levels <150 mg/dl was established as a normal value and from 150 and up to > 1000 mg/dl to consider the category in a binary variable as a high triglyceride value.

Triglycerides classification:

| Range | Category | Binary |
|---|---|---|
| < 150 mg/dL | Normal | Normal |
| 150 mg/dL - 499 mg/dL | High | High |
| 500 mg/dL - 999 mg/dL | Very High | |
| 1000 mg/dL > | Extremely High | |

For cholesterol, values were assigned based on whether patients presented three distinct categories: normal, elevated, and very high. The normal category was defined in the range < 200.0 mg/dL; for the elevated category, the ranges were established as 200.0 mg/dL - 239.9 mg/dL; finally, in the very high category, values greater than 240 mg/dL were taken.

Cholesterol classification[4]:

| Range | Category | Binary |
|---|---|---|
| < 200.0 mg/dL | Normal | Normal |
| 200.0 mg/dL - 239.9 mg/dL | High | High |
| 240.0 mg/dL > | Very High | |

[3] https://www.nhlbi.nih.gov/es/health/high-blood-triglycerides
[4] https://www.nhlbi.nih.gov/files/docs/guidelines/atglance.pdf

For creatinine, values were assigned based on the typical range for serum creatinine is:
For adult men, 0.74 to 1.35 mg/dL (65.4 to 119.3 micromoles/L) For adult women, 0.59 to 1.04 mg/dL (52.2 to 91.9 micromoles/L), In this particular case, binarization was carried out to establish normal values and high values. Taking into account the previous consideration, a cohort point was established as < 1.35 mg/dl in the case of men and < 1.04 in women to establish normal values and > 1.35 mg/dl and > 1.04 mg/dl establishing the values as high.

Creatinine classification[5]:

| Range | Category | Binary |
|---|---|---|
| For men | | |
| < 0.74 mg/dL | Low | Normal |
| 0.74 - 1.35 mg/dL | Normal | |
| 1.35 > mg/dL | High | High |
| For women | | |
| < 0.59 mg/dL | Low | Normal |
| 0.59 - 1.04 mg/dL | Normal | |
| 1.04 > mg/dL | High | High |

[5]https://www.mayoclinic.org/tests-procedures/creatinine-test/about/pac-20384646#:~:text=The%20typical%20range%20for%20serum,52.2%20to%2091.9%20micromoles%2FL

### G. Table of experiments

In our study, we employed a diverse set of artificial intelligence machinery and tools, including various machine learning models, pre-processing algorithms, filters, and other transformation steps. Here's a brief description of the algorithms used in our analysis:

**1. Machine Learning Classifiers**

- **Logistic Regression with Lasso Regularization.** Logistic Regression is a linear model used for binary classification. Lasso regularization is employed to prevent overfitting by adding a penalty term for large coefficient values, encouraging the model to select only the most important features.
- **Extreme Gradient Boosting (XGBoost).** XGBoost is an ensemble learning method that uses a collection of decision trees. It creates a strong predictive model by combining the predictions from multiple weak learners, in this case, decision trees. The algorithm is particularly powerful due to its ability to handle complex relationships in the data.
- **Multilayer Perceptron (MLP).** MLP is a type of neural network with multiple layers, including an input layer, one or more hidden layers, and an output layer. Each neuron in the input layer represents a variable in the dataset. The network learns complex patterns and relationships in the data through the hidden layers, and the output layer produces the final prediction.
- **Gaussian Naïve Bayes Classifier.** Naïve Bayes is a probabilistic classification algorithm based on Bayes' theorem. The Gaussian Naïve Bayes variant assumes that the features are normally distributed. It is particularly suited for continuous data and works well when the features are conditionally independent given the class variable.
- **Bernoulli Naïve Bayes**. Bernoulli Naïve Bayes is a variant of the Naïve Bayes algorithm specifically designed for binary feature variables. It assumes that the presence or absence of a particular feature is a binary outcome. Despite its simplicity, it performs well in classification tasks with binary or Boolean features.
- **Nearest Centroid**. Nearest Centroid is a simple and interpretable classification algorithm. It calculates the centroid (mean) of each class in the feature space and assigns new data points to the class whose centroid is closest to the point. It's particularly useful for high-dimensional data with discrete features.
- **Quadratic Discriminant Analysis (QDA)**. QDA is a variant of the Discriminant Analysis algorithm. Unlike Linear Discriminant Analysis (LDA), QDA does not assume equal covariance among classes. It estimates a separate covariance matrix for each class, allowing for more complex decision boundaries in the feature space.
- **Passive Aggressive**. Passive Aggressive is an online learning algorithm for classification and regression tasks. It is especially useful for large-scale data streams where the learning algorithm needs to adapt to new data points incrementally. The "passive-aggressive" name comes from its behavior: it makes aggressive updates for misclassified points and passive updates for correctly classified points.

- **Decision Tree**. Decision Tree is a non-parametric supervised learning method used for both classification and regression tasks. It recursively splits the dataset into subsets based on the most significant feature at each node. This process creates a tree-like model of decisions, making it easy to understand and interpret.
- **k-Nearest Neighbors Classifier (KNC)**. K-Nearest Neighbors is a non-parametric, instance-based learning algorithm. Given a new data point, KNC finds the K nearest data points in the training set and predicts the class label (for classification) or the value (for regression) based on the majority class or mean value of its neighbors.
- **Stochastic Gradient Descent Classifier (SGDC)**. This is an iterative optimization algorithm used in machine learning, especially for large datasets. Unlike traditional methods, it updates the model parameters using only one or a few training examples at a time. This approach makes it computationally efficient and well-suited for high-dimensional data. By iteratively adjusting parameters in the direction that minimizes the loss function, SGD efficiently finds optimal solutions, making it popular for real-world applications.
- **Random Forest**. Random Forest is an ensemble learning method that constructs multiple decision trees during training and outputs the class that is the mode of the classes (classification) of the individual trees. It combines the predictions of multiple decision trees to improve accuracy and generalization.
- **Extremely Randomized Trees**. Also known as Extra Trees, is another ensemble learning method that builds multiple decision trees with random splits. It further randomizes the decision tree construction by selecting random thresholds for each feature, making it even more robust to overfitting than traditional Random Forest.

**2. Class Balancing**: Class balancing techniques are used to address class imbalance in the dataset. This ensures that the machine learning models are not biased towards the majority class, leading to more accurate predictions for both classes. We used Random Under Sampling (RUS).

**3. Data standardization:** Standardization scales the features to have a mean of 0 and a standard deviation of 1. This ensures that all features contribute equally to the model and prevents features with larger scales from dominating the learning process.

- **Yeo-Johnson.** These are methods for transforming skewed data distributions into more symmetric, normal-like distributions, making the data more suitable for modeling.

**4. Feature Selection:** Feature selection techniques are applied to choose the most relevant features for the model. This reduces dimensionality and can improve the model's performance and interpretability.

- Chi-squared, using the best N=100 variables.
- Model 1: utilizes all 447 variables in the database and serves as the benchmark.
- Model 2: Experted-selected model with 13 variables (shown below).

| **Variable** | **Description** | **Category** |
|---|---|---|
| years_since_dx, | The number of years elapsed since the diagnosis of the disease (e.g., diabetes type 2 or hypertension). | Demographics |
| cs_sex | A categorical variable indicating the gender of individuals (e.g., "male" or "female"). | Demographics |
| age_at_wx_ordinal | A variable representing age in ordinal categories (e.g., "under 30 years," "30-40 years," "over 40 years"). | Demographics |
| dx_age_e11_ordinal | A variable that represents age in ordinal categories at the time of diabetes type 2 diagnosis. | Demographics |
| bmi_ordinal | A variable categorizing BMI into ranges (e.g., "underweight," "normal," "overweight," "obese"). | Measurements |
| diabetes_type_2 | A binary variable indicating whether an individual has diabetes type 2 (e.g., "yes" or "no"). | Diagnosis |
| essential_(primary)_hypertension | A binary variable indicating whether an individual has essential hypertension (e.g., "yes" or "no"). | Diagnosis |
| Dyslipidemia (E78) | A binary variable indicating whether an individual has disorders of lipoprotein metabolism and other lipidemias (e.g., "yes" or "no"). | Diagnosis |
| gfr_ordinal | A variable categorizing GFR into ranges (e.g., "normal," "mild decrease," "moderate decrease," "severe decrease"). | Laboratory |
| glucose_ordinal | A variable categorizing blood glucose levels (e.g., "normal," "prediabetes," "diabetes"). | Laboratory |
| HbA1c_ordinal | A variable categorizing hemoglobin levels into ranges (e.g., "low," "normal," "high"). | Laboratory |
| triglycerides_ordinal | A variable categorizing triglyceride levels into ranges (e.g., "low," "normal," "high"). | Laboratory |
| cholesterol_ordinal | A variable categorizing cholesterol levels into ranges (e.g., "low," "normal," "high") | Laboratory |

- Model 3: Demographics only model with 9 variables.
- Model 4: Demographics + laboratory values model with 115 variables.
- Model 5: Demographics + diagnoses values model with 163 variables.
- Model 6: Demographics + drugs values model with 119 variables.

The results for all the experiments explored are presented in the table below, marking those presented in the main manuscript in blue.

| # | Class Balancing | Data Normalization | Standardization | Feature Selection | Model Training | Nephropathy AUC |
|---|---|---|---|---|---|---|
| 1 | Yes | Yeo Johnson | Zscore | All Data | Gaussian NB | 0.856 |
| 2 | Yes | Yeo Johnson | Zscore | Expert_13 | Gaussian NB | 0.835 |
| 3 | Yes | Yeo Johnson | Zscore | Dem | Gaussian NB | 0.832 |
| 4 | Yes | Yeo Johnson | Zscore | Dem + Labs | Gaussian NB | 0.816 |
| 5 | Yes | Yeo Johnson | Zscore | Dem + Dx | Gaussian NB | 0.863 |
| 6 | Yes | Yeo Johnson | Zscore | Dem + Drugs | Gaussian NB | 0.857 |
| 7 | No | Quantile Transform | Zscore | All Data | Gaussian NB | 0.856 |
| 8 | Yes | Quantile Transform | Zscore | All Data | Gaussian NB | 0.856 |
| 9 | Yes | Yeo Johnson | Zscore | All Data | Gaussian NB | 0.855 |
| 10 | No | Yeo Johnson | Zscore | All Data | Gaussian NB | 0.854 |
| 11 | No | Quantile Transform | Zscore | All Data | Bernoulli NB | 0.818 |
| 12 | No | Yeo Johnson | Zscore | All Data | Bernoulli NB | 0.815 |
| 13 | No | Yeo Johnson | Zscore | chi2 | Quadratic Discriminant | 0.807 |
| 14 | Yes | Yeo Johnson | Zscore | All Data | Bernoulli NB | 0.780 |
| 15 | Yes | Quantile Transform | Zscore | All Data | Bernoulli NB | 0.779 |
| 16 | No | Quantile Transform | Zscore | chi2 | Bernoulli NB | 0.770 |
| 17 | Yes | Yeo Johnson | Zscore | chi2 | Bernoulli NB | 0.750 |
| 18 | Yes | Yeo Johnson | Zscore | All Data | Nearest Centroid | 0.750 |
| 19 | Yes | Quantile Transform | Zscore | chi2 | Bernoulli NB | 0.749 |
| 20 | Yes | Quantile Transform | Zscore | All Data | Nearest Centroid | 0.749 |
| 21 | No | Quantile Transform | Zscore | chi2 | Gaussian NB | 0.740 |

| | | | | | | |
|---|---|---|---|---|---|---|
| 22 | No | Yeo Johnson | Zscore | All Data | Nearest Centroid | 0.726 |
| 23 | No | Quantile Transform | Zscore | All Data | Nearest Centroid | 0.726 |
| 24 | No | Yeo Johnson | Zscore | chi2 | Bernoulli NB | 0.725 |
| 25 | Yes | Yeo Johnson | Zscore | chi2 | Nearest Centroid | 0.722 |
| 26 | No | Yeo Johnson | Zscore | chi2 | Gaussian NB | 0.722 |
| 27 | Yes | Quantile Transform | Zscore | chi2 | Nearest Centroid | 0.712 |
| 28 | Yes | Yeo Johnson | Zscore | chi2 | Gaussian NB | 0.706 |
| 29 | Yes | Quantile Transform | Zscore | chi2 | Gaussian NB | 0.705 |
| 30 | No | Quantile Transform | Zscore | chi2 | Nearest Centroid | 0.700 |
| 31 | No | Yeo Johnson | Zscore | chi2 | Nearest Centroid | 0.697 |
| 32 | Yes | Yeo Johnson | Zscore | chi2 | Quadratic Discriminant | 0.692 |
| 33 | No | Quantile Transform | Zscore | chi2 | Quadratic Discriminant | 0.680 |
| 34 | Yes | Yeo Johnson | Zscore | All Data | Quadratic Discriminant | 0.636 |
| 35 | Yes | Quantile Transform | Zscore | chi2 | Quadratic Discriminant | 0.612 |
| 36 | Yes | Quantile Transform | Zscore | chi2 | Passive Aggressive | 0.576 |
| 37 | No | Yeo Johnson | Zscore | All Data | Passive Aggressive | 0.552 |
| 38 | Yes | Yeo Johnson | Zscore | All Data | Passive Aggressive | 0.547 |
| 39 | No | Yeo Johnson | Zscore | chi2 | Passive Aggressive | 0.540 |
| 40 | Yes | Quantile Transform | Zscore | chi2 | decision_tree | 0.540 |

| | | | | | | |
|---|---|---|---|---|---|---|
| 41 | No | Quantile Transform | Zscore | All Data | decision_tree | 0.532 |
| 42 | Yes | Yeo Johnson | Zscore | All Data | decision_tree | 0.530 |
| 43 | Yes | Quantile Transform | Zscore | All Data | decision_tree | 0.529 |
| 44 | No | Yeo Johnson | Zscore | All Data | knc | 0.528 |
| 45 | No | Quantile Transform | Zscore | All Data | sgdc | 0.528 |
| 46 | Yes | Yeo Johnson | Zscore | All Data | knc | 0.527 |
| 47 | No | Yeo Johnson | Zscore | All Data | sgdc | 0.526 |
| 48 | No | Quantile Transform | Zscore | All Data | mlpc | 0.526 |
| 49 | Yes | Yeo Johnson | Zscore | chi2 | decision_tree | 0.525 |
| 50 | Yes | Yeo Johnson | Zscore | chi2 | knc | 0.525 |
| 51 | Yes | Quantile Transform | Zscore | All Data | Quadratic Discriminant | 0.524 |
| 52 | No | Yeo Johnson | Zscore | chi2 | sgdc | 0.524 |
| 53 | No | Yeo Johnson | Zscore | chi2 | knc | 0.524 |
| 54 | Yes | Quantile Transform | Zscore | chi2 | mlpc | 0.522 |
| 55 | No | Yeo Johnson | Zscore | chi2 | decision_tree | 0.522 |
| 56 | No | Yeo Johnson | Zscore | All Data | Quadratic Discriminant | 0.521 |
| 57 | Yes | Yeo Johnson | Zscore | chi2 | mlpc | 0.520 |
| 58 | No | Yeo Johnson | Zscore | All Data | mlpc | 0.520 |
| 59 | No | Yeo Johnson | Zscore | chi2 | mlpc_256 | 0.520 |
| 60 | Yes | Yeo Johnson | Zscore | All Data | mlpc | 0.520 |
| 61 | No | Quantile Transform | Zscore | chi2 | decision_tree | 0.519 |
| 62 | Yes | Quantile Transform | Zscore | chi2 | knc | 0.519 |
| 63 | Yes | Quantile Transform | Zscore | chi2 | mlpc_256 | 0.519 |

| | | | | | | |
|---|---|---|---|---|---|---|
| 64 | Yes | Yeo Johnson | Zscore | All Data | mlpc_1024 | 0.519 |
| 65 | No | Yeo Johnson | Zscore | All Data | decision_tree | 0.518 |
| 66 | Yes | Yeo Johnson | Zscore | chi2 | mlpc_256 | 0.518 |
| 67 | Yes | Yeo Johnson | Zscore | All Data | sgdc | 0.517 |
| 68 | No | Yeo Johnson | Zscore | All Data | mlpc_256 | 0.517 |
| 69 | No | Yeo Johnson | Zscore | chi2 | mlpc_1024 | 0.516 |
| 70 | No | Quantile Transform | Zscore | All Data | mlpc_256 | 0.516 |
| 71 | Yes | Yeo Johnson | Zscore | All Data | mlpc_256 | 0.515 |
| 72 | No | Quantile Transform | Zscore | chi2 | knc | 0.515 |
| 73 | No | Quantile Transform | Zscore | chi2 | mlpc_256 | 0.514 |
| 74 | No | Quantile Transform | Zscore | All Data | Quadratic Discriminant | 0.514 |
| 75 | Yes | Quantile Transform | Zscore | All Data | mlpc_1024 | 0.514 |
| 76 | No | Yeo Johnson | Zscore | All Data | mlpc_1024 | 0.514 |
| 77 | Yes | Yeo Johnson | Zscore | chi2 | mlpc_1024 | 0.514 |
| 78 | Yes | Quantile Transform | Zscore | chi2 | mlpc_1024 | 0.514 |
| 79 | Yes | Quantile Transform | Zscore | All Data | knc | 0.513 |
| 80 | Yes | Quantile Transform | Zscore | All Data | mlpc | 0.512 |
| 81 | Yes | Quantile Transform | Zscore | All Data | sgdc | 0.512 |
| 82 | No | Quantile Transform | Zscore | chi2 | mlpc_1024 | 0.511 |
| 83 | No | Quantile Transform | Zscore | chi2 | sgdc | 0.510 |
| 84 | No | Quantile Transform | Zscore | All Data | knc | 0.509 |

| | | | | | | |
|---|---|---|---|---|---|---|
| 85 | No | Quantile Transform | Zscore | All Data | mlpc_1024 | 0.508 |
| 86 | Yes | Yeo Johnson | Zscore | chi2 | Passive Aggressive | 0.507 |
| 87 | Yes | Yeo Johnson | Zscore | All Data | xgboost_1500 | 0.507 |
| 88 | No | Quantile Transform | Zscore | All Data | Passive Aggressive | 0.506 |
| 89 | Yes | Yeo Johnson | Zscore | chi2 | xgboost_1500 | 0.506 |
| 90 | Yes | Quantile Transform | Zscore | chi2 | xgboost_1500 | 0.506 |
| 91 | No | Yeo Johnson | Zscore | All Data | xgboost_1500 | 0.506 |
| 92 | Yes | Quantile Transform | Zscore | All Data | mlpc_256 | 0.505 |
| 93 | No | Quantile Transform | Zscore | All Data | xgboost_1500 | 0.504 |
| 94 | No | Quantile Transform | Zscore | All Data | xgboost | 0.504 |
| 95 | No | Yeo Johnson | Zscore | chi2 | mlpc | 0.504 |
| 96 | Yes | Quantile Transform | Zscore | All Data | logistic | 0.503 |
| 97 | Yes | Quantile Transform | Zscore | chi2 | xgboost | 0.503 |
| 98 | Yes | Quantile Transform | Zscore | All Data | xgboost | 0.503 |
| 99 | No | Yeo Johnson | Zscore | chi2 | xgboost_1500 | 0.503 |
| 100 | Yes | Quantile Transform | Zscore | All Data | xgboost_1500 | 0.503 |
| 101 | No | Quantile Transform | Zscore | All Data | logistic | 0.502 |
| 102 | Yes | Yeo Johnson | Zscore | All Data | logistic | 0.502 |
| 103 | No | Yeo Johnson | Zscore | All Data | xgboost | 0.502 |
| 104 | No | Yeo Johnson | Zscore | chi2 | xgboost | 0.502 |
| 105 | Yes | Yeo Johnson | Zscore | All Data | xgboost | 0.502 |

| | | | | | | |
|---|---|---|---|---|---|---|
| 106 | No | Quantile Transform | Zscore | chi2 | mlpc | 0.502 |
| 107 | Yes | Quantile Transform | Zscore | chi2 | logistic | 0.501 |
| 108 | No | Yeo Johnson | Zscore | chi2 | random_forest | 0.501 |
| 109 | No | Yeo Johnson | Zscore | All Data | logistic | 0.501 |
| 110 | No | Quantile Transform | Zscore | chi2 | xgboost | 0.501 |
| 111 | No | Yeo Johnson | Zscore | chi2 | extra_trees | 0.501 |
| 112 | No | Quantile Transform | Zscore | chi2 | Passive Aggressive | 0.501 |
| 113 | Yes | Yeo Johnson | Zscore | chi2 | extra_trees | 0.501 |
| 114 | No | Quantile Transform | Zscore | chi2 | xgboost_1500 | 0.501 |
| 115 | Yes | Quantile Transform | Zscore | All Data | Passive Aggressive | 0.501 |
| 116 | No | Yeo Johnson | Zscore | All Data | random_forest | 0.500 |
| 117 | No | Yeo Johnson | Zscore | chi2 | logistic | 0.500 |
| 118 | Yes | Yeo Johnson | Zscore | chi2 | logistic | 0.500 |
| 119 | No | Quantile Transform | Zscore | chi2 | logistic | 0.500 |
| 120 | Yes | Yeo Johnson | Zscore | All Data | random_forest | 0.500 |
| 121 | Yes | Quantile Transform | Zscore | All Data | random_forest | 0.500 |
| 122 | Yes | Yeo Johnson | Zscore | chi2 | sgdc | 0.500 |
| 123 | No | Quantile Transform | Zscore | All Data | random_forest | 0.500 |
| 124 | No | Quantile Transform | Zscore | chi2 | random_forest | 0.500 |
| 125 | Yes | Yeo Johnson | Zscore | chi2 | random_forest | 0.500 |
| 126 | Yes | Quantile Transform | Zscore | chi2 | random_forest | 0.500 |
| 127 | Yes | Quantile Transform | Zscore | chi2 | sgdc | 0.500 |

| | | | | | | |
|---|---|---|---|---|---|---|
| 128 | Yes | Quantile Transform | Zscore | All Data | extra_trees | 0.500 |
| 129 | No | Quantile Transform | Zscore | All Data | extra_trees | 0.500 |
| 130 | No | Yeo Johnson | Zscore | All Data | extra_trees | 0.500 |
| 131 | Yes | Yeo Johnson | Zscore | All Data | extra_trees | 0.500 |
| 132 | Yes | Yeo Johnson | Zscore | chi2 | xgboost | 0.500 |
| 133 | No | Quantile Transform | Zscore | chi2 | extra_trees | 0.500 |
| 134 | Yes | Quantile Transform | Zscore | chi2 | extra_trees | 0.500 |
